# Altered DNA methylation pattern characterizes the peripheral immune cells of patients with autoimmune hepatitis

**DOI:** 10.1101/2021.07.01.21259836

**Authors:** Kalliopi Zachou, Pinelopi Arvaniti, Aggeliki Lyberopoulou, Eirini Sevdali, Matthaios Speletas, Maria Ioannou, George K. Koukoulis, Yves Renaudineau, George N. Dalekos

## Abstract

**Objective:** AIH is a chronic liver disease of unknown aetiology with favourable response to immunosuppression. Little is known about the impact of methylation modifications on disease pathogenesis.

**Design:** 10 patients with AIH at diagnosis (time-point 1; tp1), 9 with primary biliary cholangitis (PBC) and 10 healthy controls (HC) were investigated. 8/10 AIH patients were also investigated following biochemical remission (time-point 2; tp2). Peripheral CD19(+)- and CD4(+)-cells were isolated to study global DNA methylation (5^m^C)/hydroxymethylation (5^hm^C) by ELISA and mRNA of DNA methylation (DNMT1/3A/3B)/hydroxymethylation enzymes (TET1/2/3) by quantitative RT-PCR. Epigenome wide association study (EWAS) was performed in CD4(+)-cells (Illumina HumanMethylation 850K array) in AIH and HC. Differences in total 5^m^C/5^hm^C state between AIH-tp1 and HC were also assessed by immunohistochemistry (IHC) on paraffin embedded liver sections.

**Results:** Reduced TET1 and increased DNMT3A mRNA levels characterized CD19(+) and CD4(+) lymphocytes from AIH-tp1 patients compared to HC and PBC respectively, without affecting global DNA 5^m^C/5^hm^C. In AIH-tp1, CD4(+) DNMT3A expression was negatively correlated with serum IgG (p=0.03). In remission (AIH-tp2), DNMT3A decreased in both CD19(+) and CD4(+)-cells (p=0.02, p=0.03, respectively). EWAS in CD4(+)-cells from AIH patients confirmed important modifications in genes implicated in immune responses (HLA-DP, TNF, lnRNAs and CD86). IHC confirmed increased 5^hm^C staining of periportal infiltrating lymphocytes in AIH-tp1.

**Conclusion:** Altered expression of TET1 and DNMT3A, characterizes peripheral immune cells in AIH. DNMT3A is associated with disease activity and decreased following therapeutic response. Gene specific DNA methylation modifications affect immunologic pathways that may play an important role in AIH pathogenesis.

**Summary box:** What is already known?

Autoimmune hepatitis (AIH) is a non-resolving chronic liver disease of unknown aetiology and favourable response to immunosuppression. Since the interplay between the genetic background and the environment seems to be fundamental for AIH pathogenesis, epigenetic modifications may be of particular importance.

What are the new findings?

We found characteristic alterations of DNA methylation in peripheral immune cells of AIH patients, which were associated with disease activity and modified by immunosuppressive treatment.

How might it impact on clinical practice in the foreseeable future?

These results provide the first evidence that epigenetics play a role in AIH pathogenesis, which may have therapeutic implications for the management of the disease.

## Introduction

Autoimmune hepatitis (AIH) is a chronic liver disease characterized by hypergammaglobulinaemia, autoantibodies, interface hepatitis and favourable response to immunosuppression (1–6). Although its aetiology remains unknown, the interaction between genetic and environmental factors seems fundamental in AIH pathogenesis (2,3,6).

Genetic predisposition to AIH has been linked to genes within the human leucocyte antigen (HLA) region, particularly with the allelic variants *HLA DRB1*0301* and *DRB1*0401* of DRB1, while weaker associations have been found with non-HLA genes (7,8). However, *HLA DRB1*0301* and *DRB1*0401* genotypes associations occur in only 51-55% of patients among different ethnicities, age groups and geographic regions, indicating that additional factors, such as epigenetic changes, could contribute to its pathogenesis (7,9).

Epigenetic modifications, consisting of DNA methylation, histone adjustment and micro-RNAs (miRNAs), influence gene expression without altering the DNA sequence. In eukaryotic cells, DNA methylation represents the central epigenetic process and refers to the methylation of carbon 5 of cytosine (5^m^C) at the cytosine-phosphate-guanine dinucleotides (CpGs) by DNA methyl-transferases (DNMT1,3A,3B) (10-12). The latter is counterbalanced by an active DNA demethylation process comprising of the oxidization of 5^m^C into 5-hydroxymethylcytosine (5^hm^C) by Ten Eleven Translocation (TET1,2,3) deoxygenases (13).

Epigenetic studies have been performed mainly in liver fibrosis, non-alcoholic fatty liver disease, hepatocellular carcinoma and cholangiocarcinoma (14,15). To date, only few studies investigated the epigenetic changes in autoimmune liver diseases. In primary biliary cholangitis (PBC), hypomethylation of several gene promoters, especially within the X chromosome, has been documented (16-18). Additionally, different expressions of several miRNAs have been reported in PBC and primary sclerosing cholangitis (19-22). In AIH, epigenetic data is even more limited, with only one study reporting significant association of elevated miR21 with the biochemical and histological activity of AIH and decreased miR21 and miR122 in cirrhotic patients (23).

As from the best of our knowledge, DNA 5^m^C/5^hm^C have not been explored in AIH, we investigated the potential presence of DNA methylation modifications in peripheral B- and T-cells from AIH patients by assessing alterations in DNA methylation through the analysis of global 5^m^C and DNMT1/3A/3B expression as well as alteration in 5^hm^C and TETs transcriptional expression levels. Such analysis was performed at diagnosis and remission. Differences in total 5^m^C/5^hm^C state between AIH patients at diagnosis and healthy controls (HC) were also assessed by immunohistochemistry (IHC) on paraffin embedded liver sections. Finally, in an attempt to evaluate the methylation alterations in specific CpG sites across the whole genome, we performed epigenome wide association study (EWAS) in CD4(+)-cells from patients and controls using the Illumina HumanMethylation 850K array.

## Patients and Methods

### Study samples and peripheral mononuclear cells

*(PBMCs) preparation* PBMCs from 10 AIH patients were isolated from peripheral blood collected at the time of diagnosis (AIH time-point 1; AIH-tp1), by gradient centrifugation (Histopaque-1077, Sigma-Aldrich, St. Louis, MI, USA). PBMCs were then mixed with freezing medium (FBS with 10% DMSO Sigma-Aldrich) and cryopreserved in liquid nitrogen until use.

In eight of these patients, PBMCs were also isolated at a second time-point when they had complete biochemical response under immunosuppression (AIH time-point 2; AIH-tp2). According to the guidelines of the Hellenic Association for the Study of the Liver (5) and our published protocols (24,25), AIH-tp2 patients were receiving at the time of investigation either combination therapy with prednisolone 0.5-1mg/kg/day and mycophenolate mofetil 1.5-2g/day (MMF; n=6) or MMF maintenance monotherapy (n=2). Ten healthy served as HC and 9 PBC patients at diagnosis before treatment initiation served as the disease control group. Patients and controls were age- and sex-matched (Supplementary Table 1).

As in our previous reports (24,25), all biopsies were assessed using the Knodell histologic/activity index score (26) and patients were divided into two groups according to inflammation: minimal-mild and moderate-severe and according to fibrosis: minimal/mild-moderate and severe fibrosis-cirrhosis. In PBC, the Ludwig staging system was applied (27).

All patients consented to participate in this study. The ethical committee of the General University Hospital of Larissa approved the protocol which conforms to the ethical guidelines of the 1975 Declaration of Helsinki as reflected in a priori approval by the institution’s human research committee (21-03-2016/2258).

### CD19(+) B- and CD4(+) T-cells isolation

CD19(+) and CD4(+)-lymphocytes were isolated by ROBOSEPTM-S platform (Stemcell Technologies, Vancouver, USA). CD19(+) B- and CD4(+) T-cells were incubated with specific antibodies for magnetic selection using the EasySep Human CD19 Positive Selection Kit II and Human CD4 Negative Selection Kit, respectively (Stemcell Technologies). Purity of the isolated B- and T-cells was assessed by flow cytometry using PE-Cy5-anti-CD19 and FITC-anti-CD3/PE-anti-CD8 antibody (Biolegend Inc., San Diego, CA, USA) on a Coulter FC-500 flow cytometer (Beckman-Coulter, Brea, CA, USA) (Supplementary Figures 1A-1D). A total average of 1×10^6^ CD19(+) and 2×10^6^ CD4(+)-cells were isolated per sample and stored at -80^0^ C.

### DNA/RNA extraction and quantification

Genomic DNA was extracted from CD19(+) and CD4(+)-cells with QIAamp Blood mini purification kit (Qiagen, Hilden, Germany). Quantification was measured at 260nm in a UV-VIS Nanodrop spectrophotometer (Thermo Fisher Scientific, Waltham, MA, USA). DNA purity was determined by the ratio of 260nm to 280nm absorbance levels. Approximately, 0.5-1μg of genomic DNA was isolated from CD19(+) and 1-3μg from CD4(+)-cells.

Total RNA was isolated using the RNeasy mini kit (Qiagen) and quantification and purity were determined as described above. Approximately 0.5-1.2μg of total RNA was isolated from CD19 (+) and 1-3.2μg from CD4(+)-cells.

### Determination of 5^m^C and 5^hm^C DNA levels

5^m^C and 5^hm^C DNA levels were determined using MethylFlashTM Global DNA Methylation (5-mC) and Hydroxymethylation (5-hmC) ELISA Easy Kit (EpiGentek, NY, USA), according to the manufacturer’s instructions (Supplementary Patients and Methods).

### DNMT1, DNMT3A, DNMT3B, TET1, TET2 and TET3 mRNA quantification

One μg of total RNA was reversed transcribed using random hexamer primers and 10 Units of Transcriptor Reverse Transcriptase according to the Transcriptor First Strand cDNA synthesis kit (Roche Diagnostics, Basel, Switzerland) in a 20μl reaction for 10min at 25°C, 60min at 50°C and 5min at 85°C. Quantitative polymerase chain reaction was carried out using FastStart DNA Master SYBR Green I (Roche Diagnostics) in a total volume of 20μl containing 250nM specific forward and reverse primers. Amplification and detection were performed in a Lightcycler^R^ 96 Instrument (Roche Life Sciences, Bavaria, Germany) under the following conditions: 1 cycle at 50°C for 2min and 95°C for 10min and 40 cycles at 95°C for 15sec and 60°C for 1min. The primers used for DNMT3A, DNMT3B, TET1 and TET2 were as previously described (28). For the quantification of human DNMT1, TET3 and Glyceraldehyde-3-Phosphate Dehydrogenase (GAPDH) genes, commercially specific primers were used: DNMT1: Hs00945875_m1, TET3: Hs00896441_m1, GAPDH: Hs02758991_g1 (Thermo Fisher Scientific, Waltham, MA, USA). Human GAPDH mRNA was chosen as internal control and for quantification of gene expression the comparative CT method [2-(ΔCT target -ΔCT calibrator) or 2-ΔΔCT] was used. Validation experiments were carried out in duplicates and each run was completed with a melting curve analysis to confirm the specificity of the amplification and the lack of primer dimmers.

### EWAS

Five hundred ng of DNA from CD4(+)-lymphocytes from AIH (10 tp1 and 5 tp2) and 9 HC were bisulfite-converted (Zymo Research, Irvine, California, USA) and DNA methylation was evaluated by hybridising bisulfite-converted DNA to the Human Methylation EPIC array Bead Chip (Diagenode SA, Belgium), which allows the interrogation of over 850000 methylation sites throughout the genome at single-nucleotide resolution (Supplementary Patients and Methods; EWAS). These steps were performed by NXT-DX Company (Gent, Belgium) according to manufacturer’s instructions.

Methylation data was provided as β-values: β=M/(M+U), where M was the fluorescent signal of methylation and U the respective signal of the unmethylated probe. The β-values ranged from 0 (no methylation) to 1 (100% methylation). A quality control on the output of the Illumina Infinium EPIC array was performed with the Bioconductor R package, Chip Analysis Methylation Pipeline (ChAMP), according to which no sample had a proportion of failed probes *>*0.1. After normalisation, potential batch effects were evaluated with the singular value decomposition method, which did not identify any significant source of variations that needed corrections.

### Liver immunohistochemistry (IHC)

Paraffin embedded sections from 6 AIH-tp1 patients and 7 HC obtained during cholecystectomy, were investigated. Staining for 5^m^C and 5^hm^C was performed with anti-5-methylcytosine (5-mC) and anti-5-hydroxymethylcytosine (5-hmC) antibodies [Abcam, 33D3 (ab10805) and RM236 (ab214728)]. 5μm liver sections were deparaffinized by immersing the slides in xylene and in concentration decreasing alcohol grades solution. Antigen retrieval was performed in a Tris-EDTA (pH=9) solution at 98°C for 20 min, while endogenous peroxidase activity was blocked by quenching the tissue sections with 3.0% hydrogen peroxide in methanol for 10 minutes. The sections were then washed with 1% donkey serum in PBS-0,4% Triton X-100 (TBST) solution for 5 minutes for permeabilization, followed by incubation with primary antibody (dilution 1/150) in room temperature for 30 minutes. After washing the sections with TBST solution, they were incubated with Linker and Polymer HRP (Ready to use reagent, Envision Flex, Dako) for 15 and 30 minutes, respectively. Finally, sections were incubated with 3,3’-diaminobenzidin (DAB^+^) and washed with distilled water. Hematoxylin Harris was used for nuclear counterstaining and dehydration was performed in increasing concentration alcohol solutions and xylene. Negative controls consisted of substitution of primary antibody with pre-immune serum.

### Immunohistochemical evaluation

Immunostaining was semi-quantitatively evaluated in a blinded fashion regarding any of the histological and clinical characteristics of the patients by two independent observers. The degree of staining was determined according to its amount and intensity, using a 4-point scoring system, as follows: 0=no staining; 1=positive nuclear staining in less than 20% of cells; 2=21-50% of positive cells; and 3=positive nuclear staining in more than 50% of cells.

### Statistical analysis

Analysis was made using the SPSS 20 and GraphPad-Prism 7.0 software. Results were expressed as median (range) and mean±standard deviation. Data were compared with Kruskall-Wallis and Mann-Whitney U-test for the detection of differences between independent samples and Wilcoxon test for paired samples. Pearson coefficient (R) and Spearman’s coefficient (r) were used for correlations, where applicable. Two-sided *p-*values <0.05 were considered as statistically significant in 95% confidence interval.

Statistical analysis of the EWAS data was performed with the Bioconductor R package ChAMP (29). After normalization, identification of differentially methylated positions (DMPs) between groups was performed using Benjamini-Hochberg adjusted *p-*value <0.05. Differentially methylated regions (DMRs) between groups, were identified with BumpHunter method and adjusted *p*-value <0.05 (30). Gene set enrichment analysis using GOmeth method was performed on the genes associated with the DMRs to determine potential enriched pathways (31). Finally, in order to identify if differentially methylated genes shared common functional properties between AIH-tp2 and AIH-tp1, we used the Database for Annotation, Visualization and Integrated Discovery (DAVID), which builds clusters of genes with significantly similar ontologies as tested against a complete list of genes in the database (32). Medium stringency was used to yield a broader set of ontological groups.

## Results

### Global 5^m^C and 5^hm^C in CD19(+) and CD4(+)-cells and their association with clinical, biochemical and serological parameters

Global 5^m^C and 5^hm^C levels in CD4(+) and CD19(+)-cells did not differ between the study groups (Figure 1). In addition, 5^m^C and 5^hm^C levels in AIH-tp1 and AIH-tp2 patients were not correlated with age, AST, ALT, IgG, ANA and SMA (data not shown). However, 5^m^C levels in CD4(+)-lymphocytes were negatively correlated with disease duration (r=-0.76; p=0.01) in AIH-tp1 patients.

**Figure 1:**
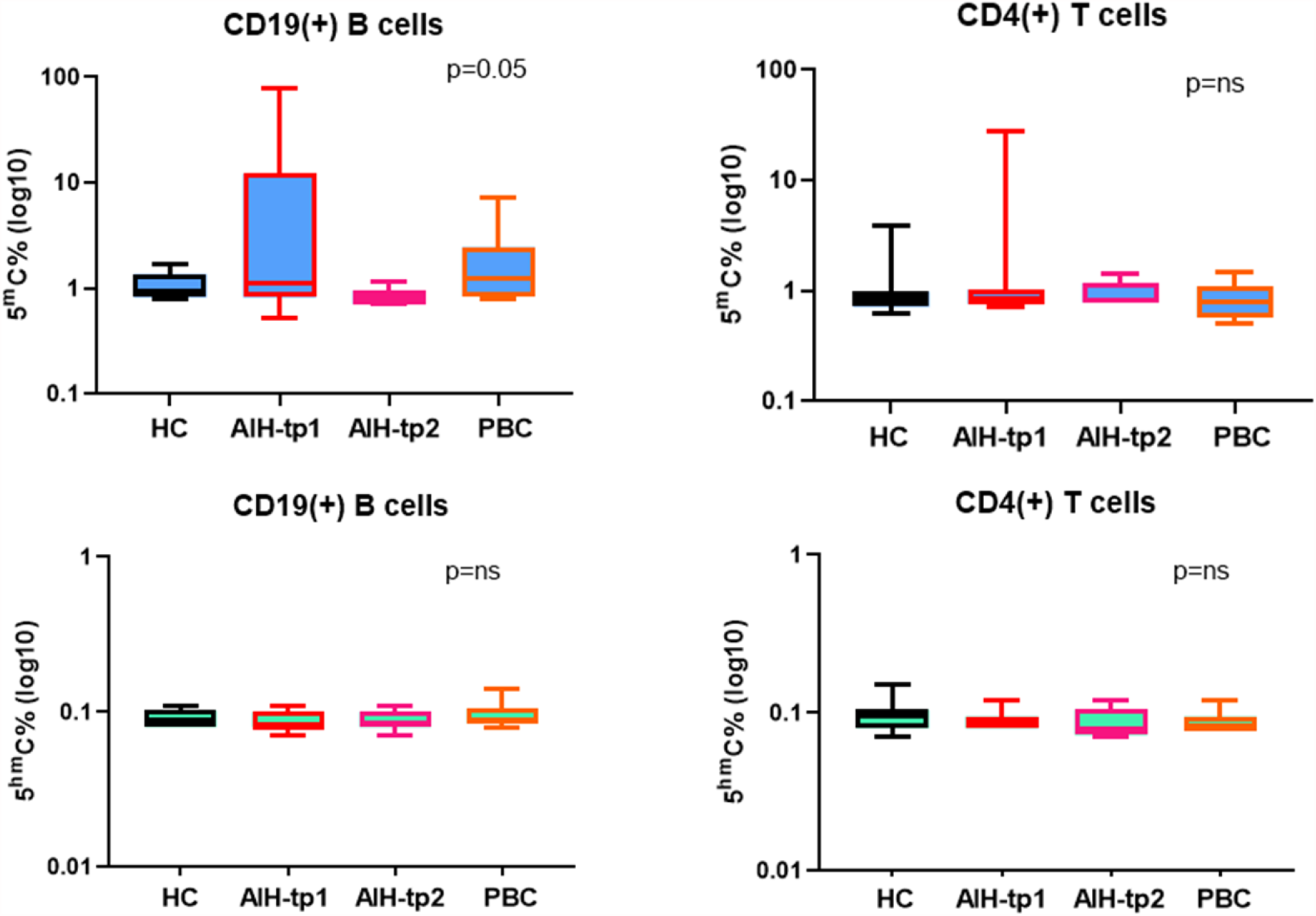
No differences were found in 5^m^C **(A and B)** and 5^hm^C levels **(C and D)** in CD19(+) and CD4(+)-cells from AIH-tp1, AIH-tp2, PBC and HC. 5^m^C, 5-methylcytosine; 5^hm^C, 5-hydroxymethylcytosine; AIH-tp1, autoimmune hepatitis time-point 1; AIH-tp2, autoimmune hepatitis time-point 2; PBC, primary biliary cholangitis; HC, healthy controls; ns, not-significant.

### DNMTs and TETs in CD19(+) and CD4(+)-lymphocytes from AIH-tp1 patients

AIH-tp1 patients had significantly lower TET1 mRNA levels in CD19(+) and CD4(+) lymphocytes compared to HC (p=0.03; p=0.01, respectively; Figures 2A, 2B). AIH-tp1 had significantly higher DNMT3A in CD19(+) and CD4(+) lymphocytes compared to PBC (p=0.04; p=0.002, respectively; Figures 2C,2D). No differences of DNMT1, TET2 and TET3 in CD19(+) or CD4(+)-cells were observed between AIH-tp1 and HC or PBC. Of note, PBC had significantly higher DNMT1 and TET3 in CD19(+)-cells compared to HC (p=0.02; p=0.005, respectively; Supplementary Figures 2A,2B). DNMT3B levels were beyond the detection threshold so no further analysis was made.

**Figure 2:**
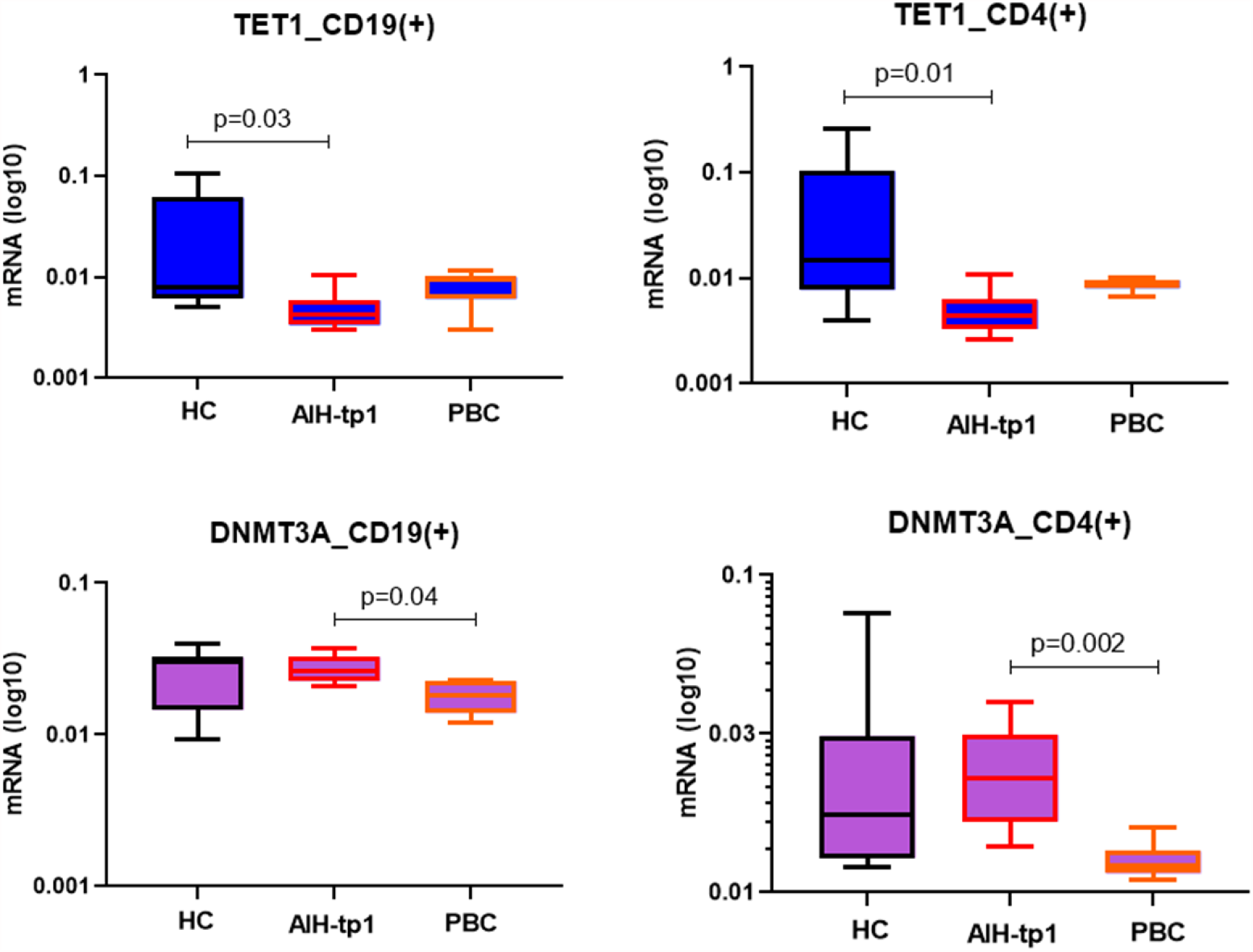
**A)** CD19(+)-cells from AIH-tp1 characterized by reduced TET1 compared to HC (p=0.03). **B)** CD4(+)-cells from AIH-tp1 characterized by reduced TET1 compared to HC (p=0.01). **C)** Increased DNMT3A was found in CD19(+)-cells from AIH-tp1 compared to PBC (p=0.04). **D)** Increased DNMT3A was found in CD4(+)-cells from AIH-tp1 compared to PBC (p=0.002). TET1, Ten-eleven translocation methylcytosine dioxygenase 1; DNMT3A, DNA methyltransferase 3A; AIH-tp1, autoimmune hepatitis time-point 1; AIH-tp2, autoimmune hepatitis time-point 2; PBC, primary biliary cholangitis; HC, healthy controls.

### Effect of immunosuppression on DNMTs and TETs expression in AIH

AIH-tp2 patients had significantly lower DNMT3A mRNA in CD19(+) (p=0.02) and CD4(+)-lymphocytes (p=0.03) compared to AIH-tp1 (Figures 3A,3B). This supports that immunosuppression controls DNMT3A overexpression as DNMT3A levels in AIH-tp2 did not differ from HC, either in CD19(+) or in CD4(+) lymphocytes (Supplementary Figure 3). The mRNA levels of DNMT1 and TETs did not differ between the two groups (data not shown).

**Figure 3:**
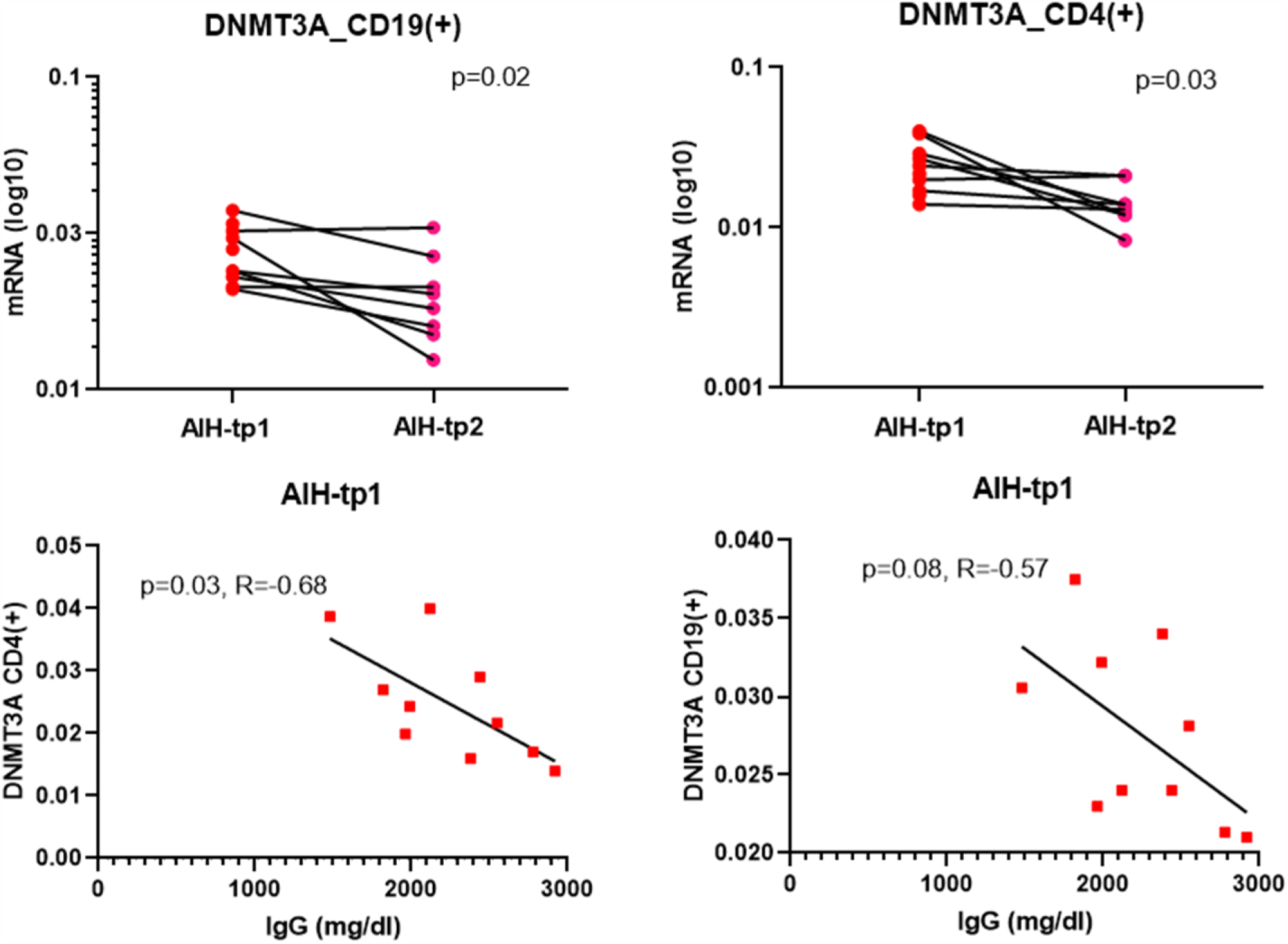
CD19(+) and CD4(+)-cells after complete remission (AIH-tp2) characterized by decreased DNMT3A mRNA levels (logarithmic scale) compared to AIH-tp1 (**A**; p=0.02 and **B**; p=0.03). **C)** DNMT3A in CD4(+)-cells was negatively correlated with IgG in AIH-tp1 (R=-0.68, p=0.03). **D)** A negative trend was observed between DNMT3A and IgG in CD19(+)-cells from AIH-tp1 (R=-0,57, p=0.08). DNMT3A, DNA methyltransferase 3A; AIH-tp1, autoimmune hepatitis time-point 1; AIH-tp2, autoimmune hepatitis time-point 2.

### Association of DNMTs and TETs mRNA with clinical, biochemical and serological parameters in AIH-tp1 and AIH-tp2 patients

DNMT3A mRNA in CD4(+)- and CD19(+)-cells of AIH-tp1 was negatively correlated with IgG (R=-0.68; p=0.03 and R=-0.57; p=0.08, respectively) (Figures 3C,3D). In contrast, DNMT3A levels in CD4(+)-cells from AIH-tp2 were positively correlated with IgG (r=0.8; p=0.02).

### EWAS in CD4(+) lymphocytes from AIH-tp1 patients compared to HC

As DNMT3A and TET1 mRNA variations were more important in CD4(+) as compared to CD19(+) lymphocytes, CD(4+)-cells were further selected to investigate DMRs, which are more highly associated with disease as compared to a single CpG analysis. To this end we have used the BumpHunter method and have identified 287 CpG motifs corresponding to 29 DMRs between 10 AIH-tp1 and 9 HC (Figure 4A). These DMRs corresponded to unique and annotated genes present on 26 autosomes and 3 on sex chromosomes. Regarding functional genomic distribution, the majority of DMRs (14/29; 48.3%), corresponded to gene promoters and transcription start sites (TSS), while 6/29 (20.7%) corresponded to introns (Figure 4B). Regarding methylation status, 17/29 (58.6%) genes were hypomethylated (Table 1). Interestingly, 7/8 differentially methylated genes located on chromosome 6 are located on the major histocompatibility complex (MHC). Among them, two are encoded by MHC class-II molecules: HLA-DPA1 (hypermethylated, p=0.003.) and HLA-DPB2 (hypomethylated, p=0.01). In addition, the long intergenic non-protein coding RNA 2571 gene (LINC02571), which belongs to the group of long noncoding RNAs (lncRNAs), was retrieved hypermethylated (p=0.02), while the promoter of the tumor necrosis factor (TNF) gene was hypomethylated (p=0.01; Table 1) and ZFP57 was hypermethylated (p=0.02).

**Figure 4:**
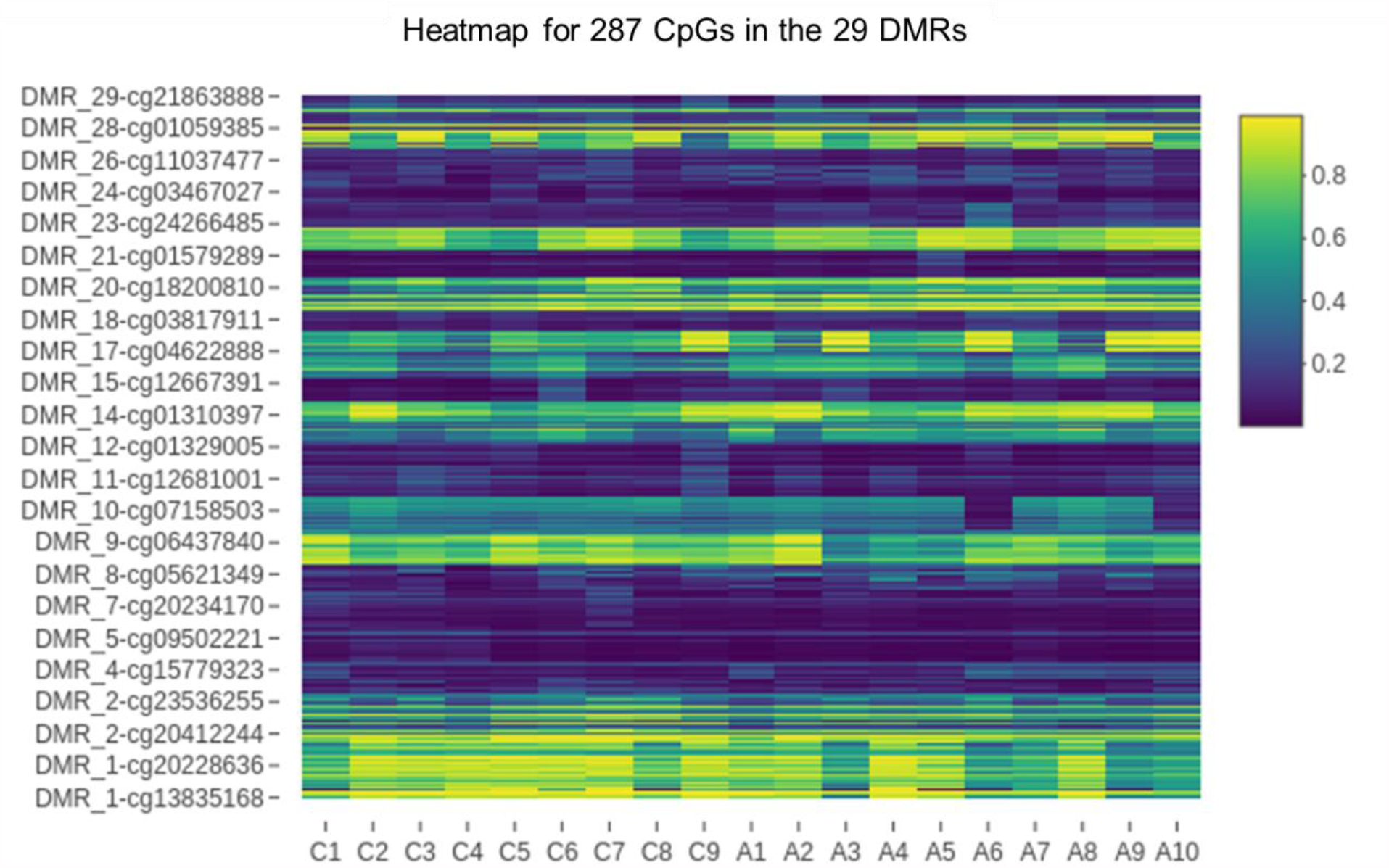
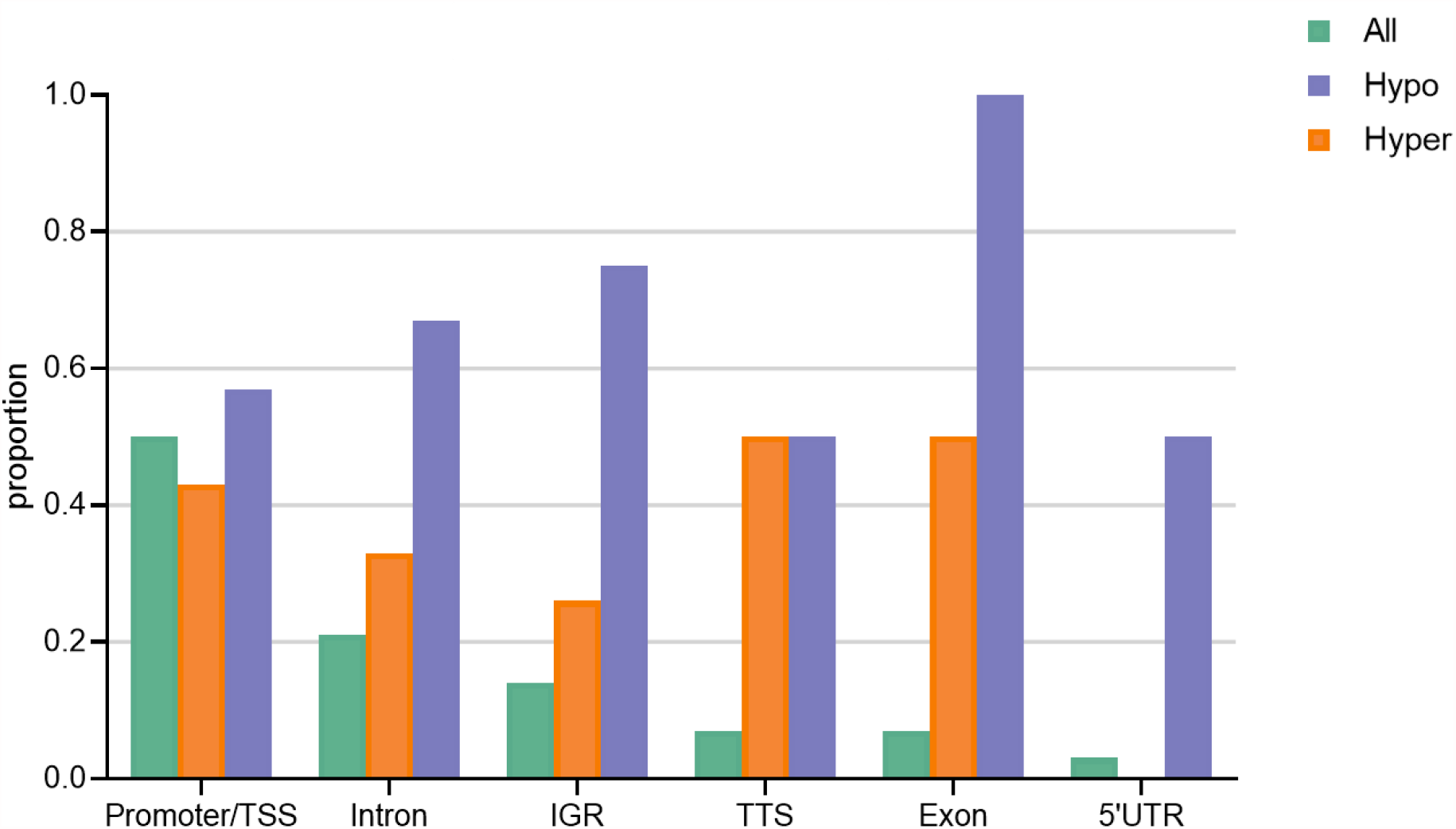
**A)** Heatmap of significant CpGs corresponding to 29 DMRs detected between AIH-tp1 and HC (C1-C9: HC, A1-A10: AIH-tp1). **B)** Genomic distribution of 29 DMRs. AIH-tp1, autoimmune hepatitis time-point 1; CpGs, cytosine-phosphate-guanine dinucleotides; DMRs, differentially methylated regions; TSS: transcription start sites, IGR: intergenic regions, 5΄UTR: 5΄untranslated region; Hypo, hypomethylated; Hyper, hypermethylated.

**Table 1:**
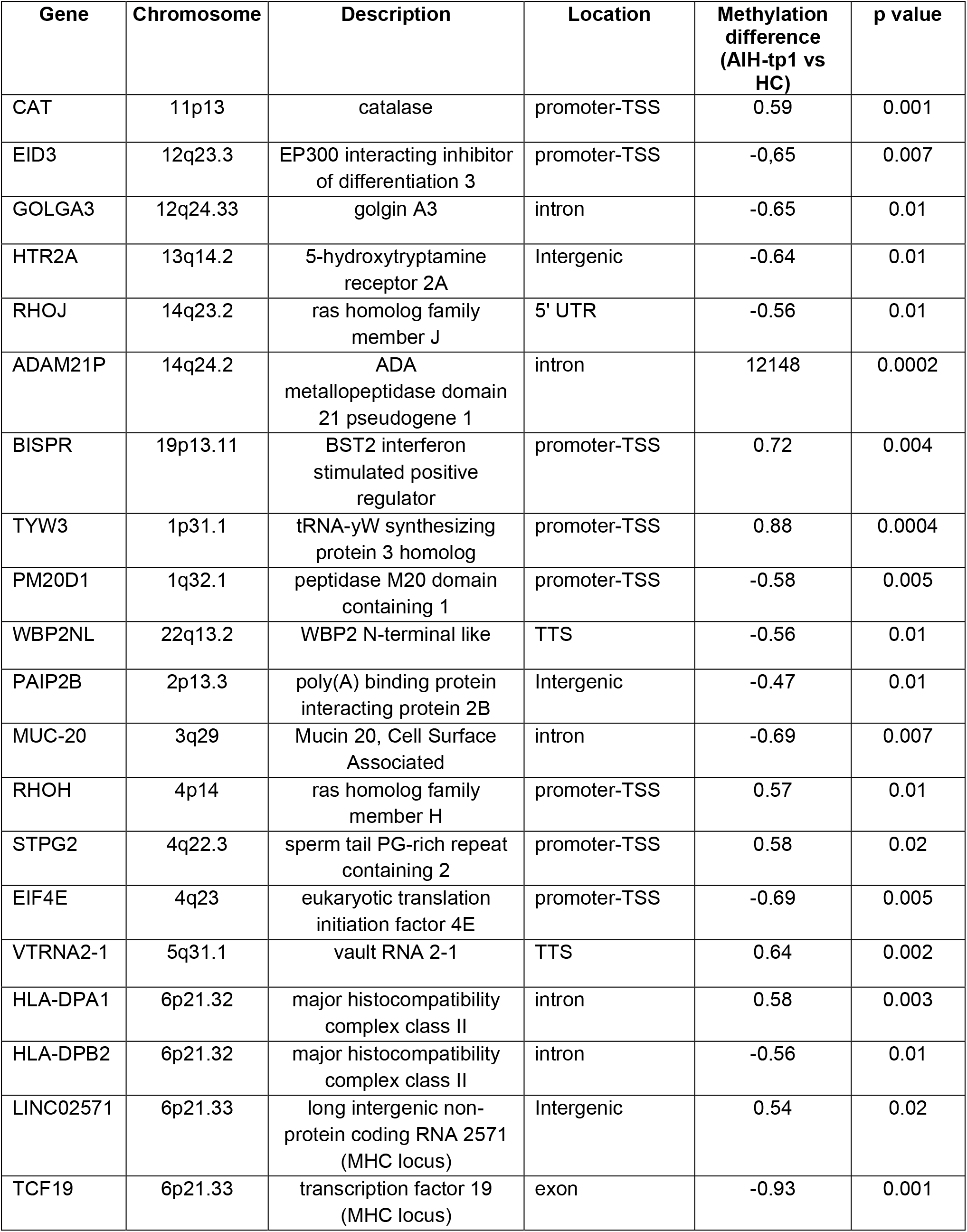
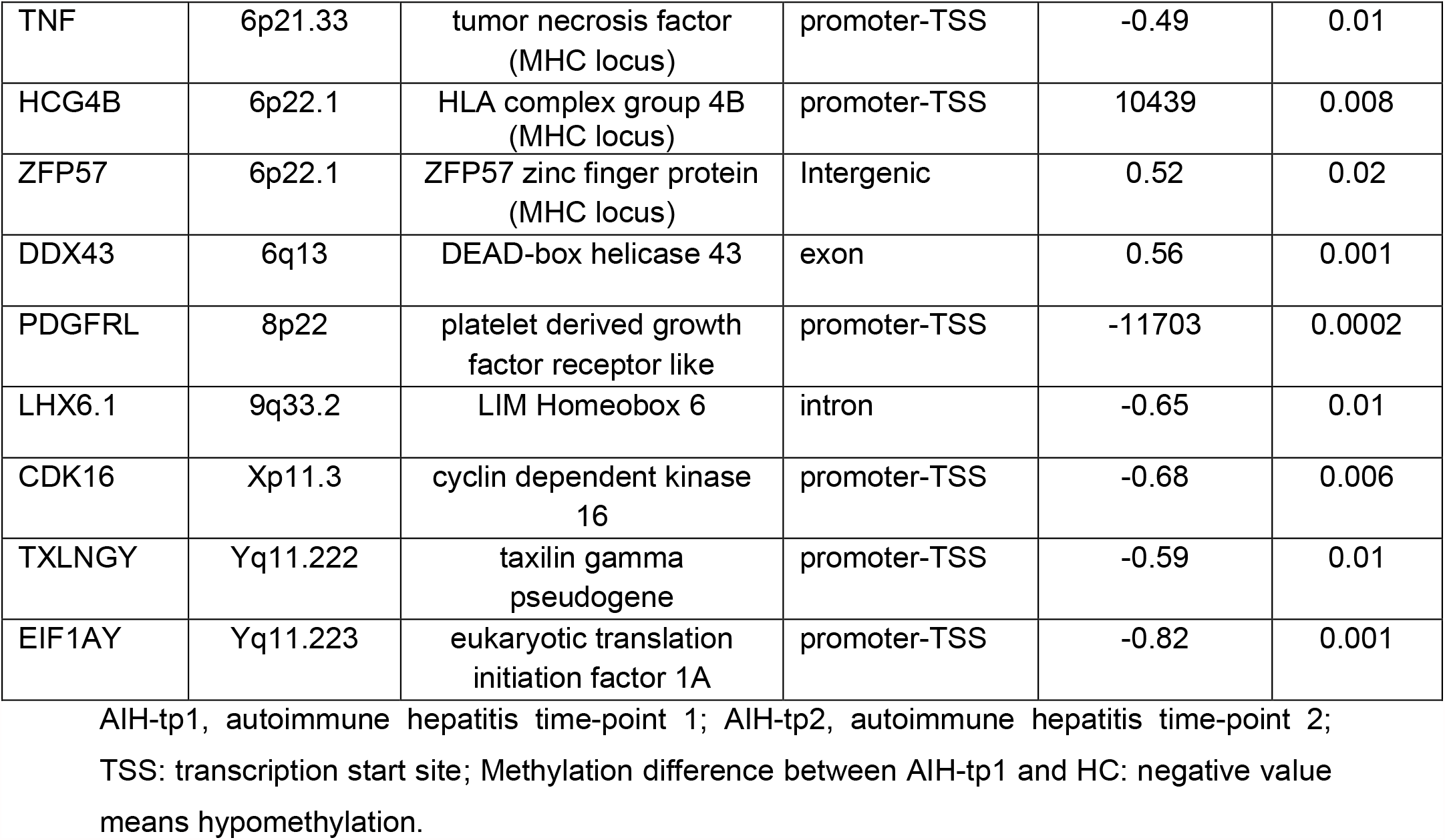
Differentially methylated genes corresponding to 29 DMRs detected in AIH-tp1 patients and healthy controls (HC).

| Gene | Chromosome | Description | Location | Methylation difference (AIH-tp1 vs HC) | p value |
| --- | --- | --- | --- | --- | --- |
| CAT | 11p13 | catalase | promoter-TSS | 0.59 | 0.001 |
| EID3 | 12q23.3 | EP300 interacting inhibitor of differentiation 3 | promoter-TSS | -0.65 | 0.007 |
| GOLGA3 | 12q24.33 | golgin A3 | intron | -0.65 | 0.01 |
| HTR2A | 13q14.2 | 5-hydroxytryptamine receptor 2A | Intergenic | -0.64 | 0.01 |
| RHOJ | 14q23.2 | ras homolog family member J | 5' UTR | -0.56 | 0.01 |
| ADAM21P | 14q24.2 | ADA metalloproteinase domain 21 pseudogene 1 | intron | 12148 | 0.0002 |
| BISPR | 19p13.11 | BST2 interferon stimulated positive regulator | promoter-TSS | 0.72 | 0.004 |
| TYW3 | 1p31.1 | tRNA-yW synthesizing protein 3 homolog | promoter-TSS | 0.88 | 0.0004 |
| PM20D1 | 1q32.1 | peptidase M20 domain containing 1 | promoter-TSS | -0.58 | 0.005 |
| WBP2NL | 22q13.2 | WBP2 N-terminal like | TTS | -0.56 | 0.01 |
| PAIP2B | 2p13.3 | poly(A) binding protein interacting protein 2B | Intergenic | -0.47 | 0.01 |
| MUC-20 | 3q29 | Mucin 20, Cell Surface Associated | intron | -0.69 | 0.007 |
| RHOH | 4p14 | ras homolog family member H | promoter-TSS | 0.57 | 0.01 |
| STPG2 | 4q22.3 | sperm tail PG-rich repeat containing 2 | promoter-TSS | 0.58 | 0.02 |
| EIF4E | 4q23 | eukaryotic translation initiation factor 4E | promoter-TSS | -0.69 | 0.005 |
| VTRNA2-1 | 5q31.1 | vault RNA 2-1 | TTS | 0.64 | 0.002 |
| HLA-DPA1 | 6p21.32 | major histocompatibility complex class II | intron | 0.58 | 0.003 |
| HLA-DPB2 | 6p21.32 | major histocompatibility complex class II | intron | -0.56 | 0.01 |
| LINC02571 | 6p21.33 | long intergenic non-protein coding RNA 2571 (MHC locus) | Intergenic | 0.54 | 0.02 |
| TCF19 | 6p21.33 | transcription factor 19 (MHC locus) | exon | -0.93 | 0.001 |
| TNF | 6p21.33 | tumor necrosis factor<br>(MHC locus) | promoter-TSS | -0.49 | 0.01 |
| HCG4B | 6p22.1 | HLA complex group 4B<br>(MHC locus) | promoter-TSS | 10439 | 0.008 |
| ZFP57 | 6p22.1 | ZFP57 zinc finger protein<br>(MHC locus) | Intergenic | 0.52 | 0.02 |
| DDX43 | 6q13 | DEAD-box helicase 43 | exon | 0.56 | 0.001 |
| PDGFRL | 8p22 | platelet derived growth<br>factor receptor like | promoter-TSS | -11703 | 0.0002 |
| LHX6.1 | 9q33.2 | LIM Homeobox 6 | intron | -0.65 | 0.01 |
| CDK16 | Xp11.3 | cyclin dependent kinase<br>16 | promoter-TSS | -0.68 | 0.006 |
| TXLNGY | Yq11.222 | taxilin gamma<br>pseudogene | promoter-TSS | -0.59 | 0.01 |
| EIF1AY | Yq11.223 | eukaryotic translation<br>initiation factor 1A | promoter-TSS | -0.82 | 0.001 |

AIH-tp1, autoimmune hepatitis time-point 1; AIH-tp2, autoimmune hepatitis time-point 2; TSS: transcription start site; Methylation difference between AIH-tp1 and HC: negative value means hypomethylation.

One of the most differentially methylated genes (p=0.0002) was the promoter of platelet derived growth factor receptor like gene. Finally, the RhoH gene was found hypermethylated (p=0.01; Table 1).

Using the GOmeth method, no pathways were identified as enriched based on the genes associated with the identified DMR.

### EWAS in CD4(+) lymphocytes from AIH-tp1 patients compared to AIH-tp2

Next, we investigated whether methylation of the CpG motifs is affected by immunosupression in CD4(+) lymphocytes from AIH (10 AIH-tp1 and 5 AIH-tp2). In AIH-tp2 and at CpG level, 831 differentially methylated probes (DMPs; 11,2% hypomethylated and 88,2% hypermethylated) were identified compared to AIH-tp1, corresponding to 576 unique and annotated genes (Figure 5A). DMPs functional genomic distribution retrieved an enrichment in intergenic regions at isolated CpG or open sea regions as well as in gene bodies within shores regions present up to 2kb from CpG islands (Figures 5B,5C).

**Figure 5:**
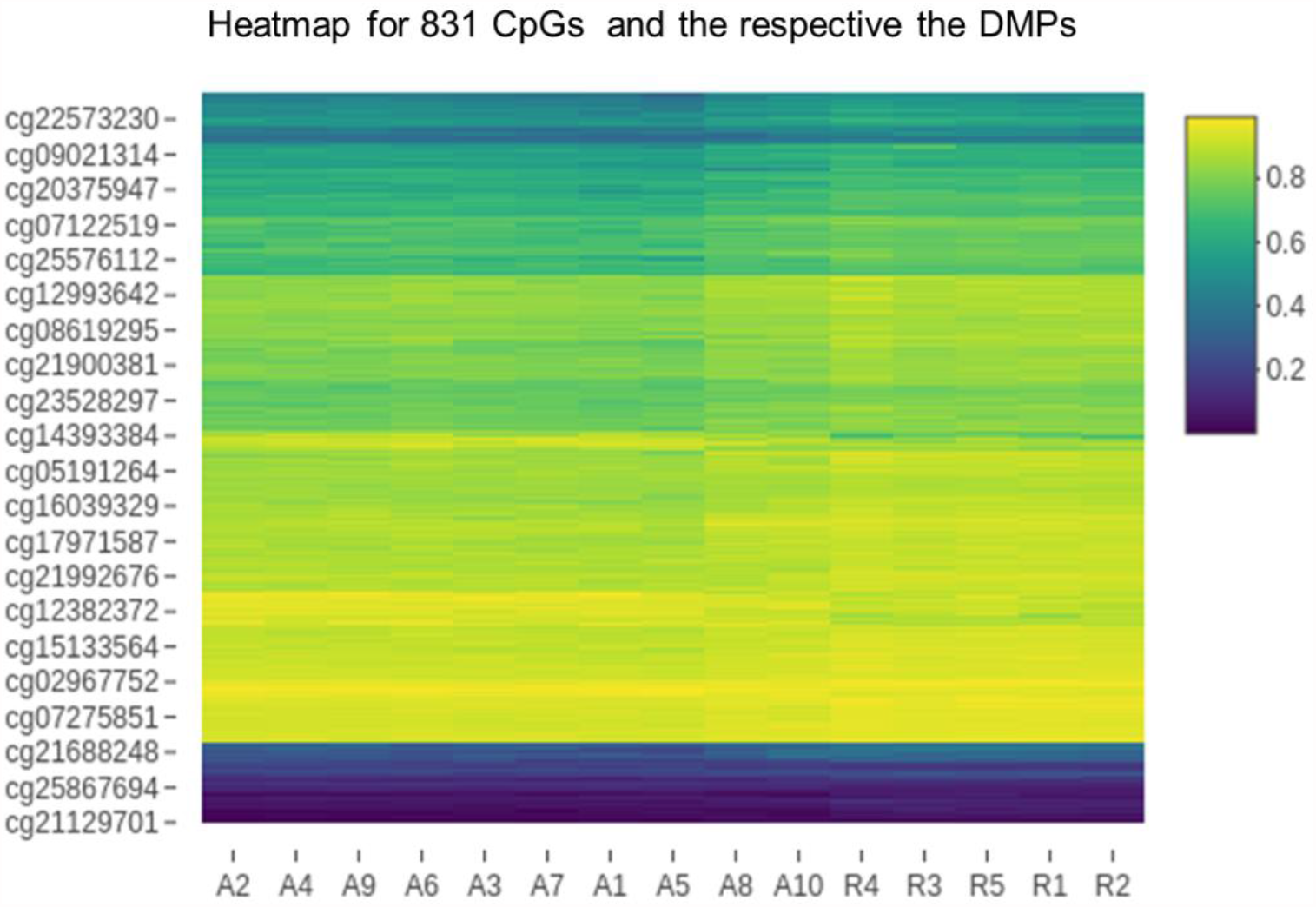
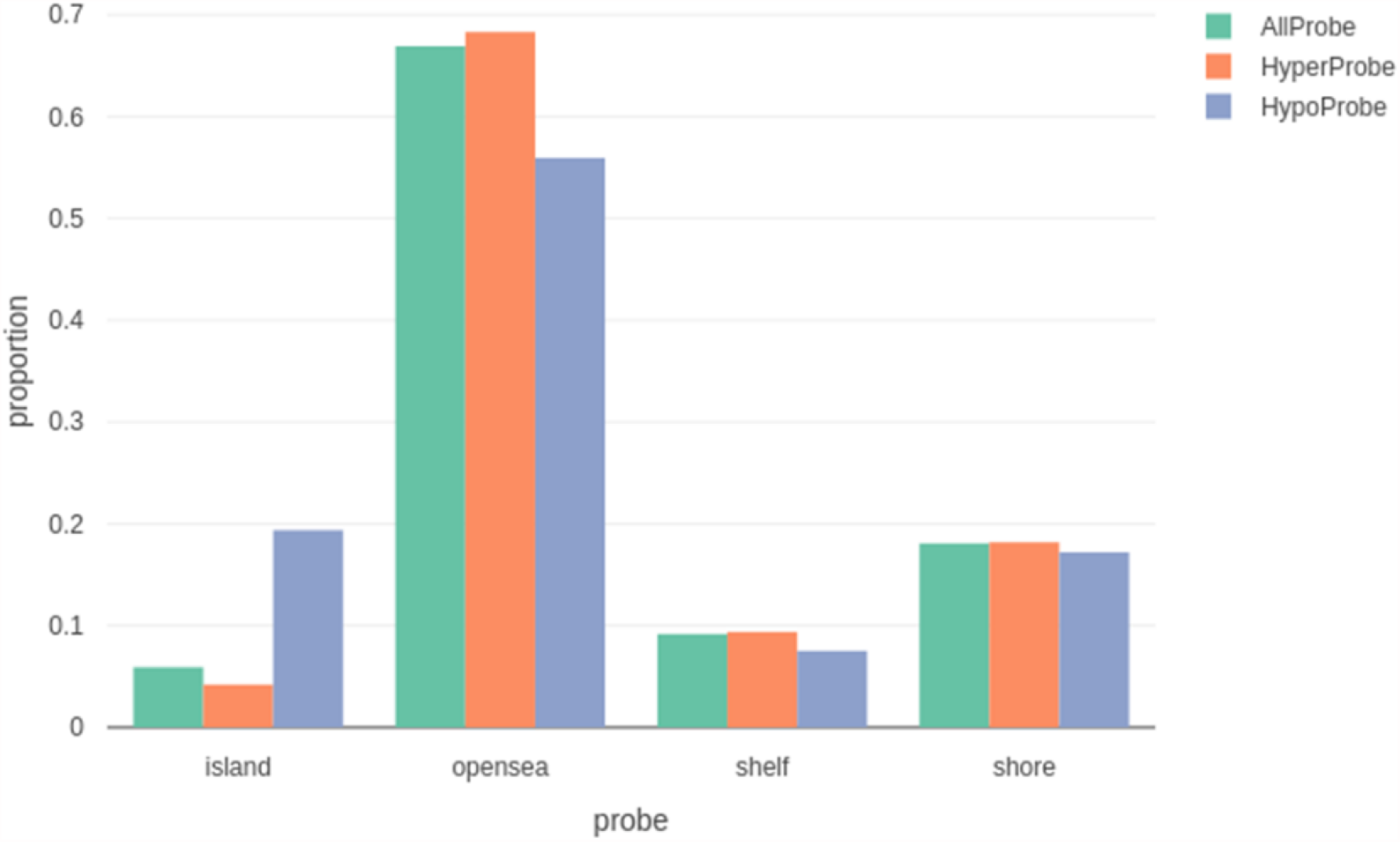
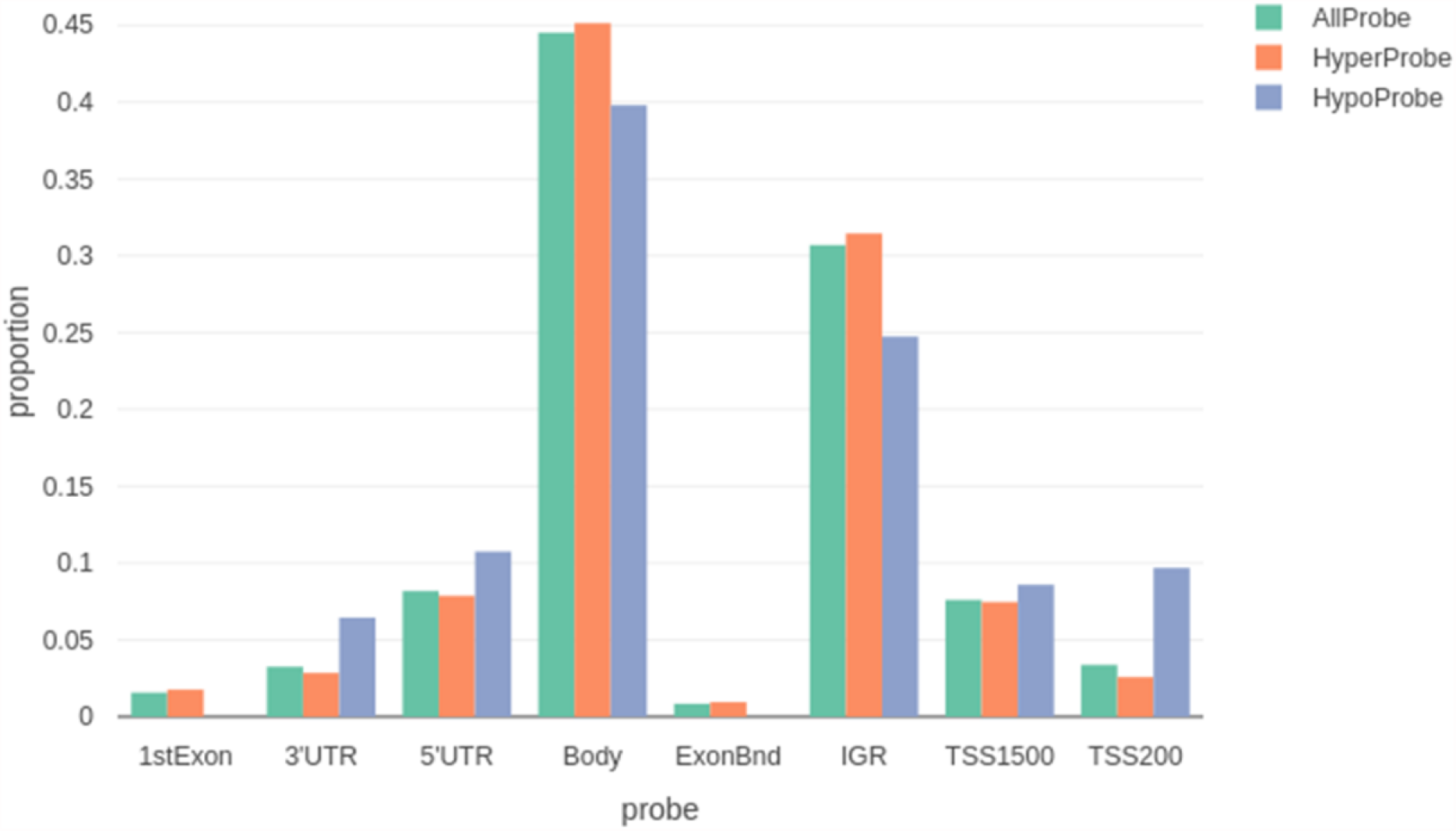
**A)** Heatmap of significant CpGs corresponding to 831 DMPs detected between AIH-tp2 (n=5) and AIH-tp1 (n=10) (R1-5: AIH-tp2, A1-10: AIH-tp1). **B)** Genomic distribution of significant DMPs. DMPs are enriched in open sea regions and shores. **C)** Most DMPs are located on gene bodies and IGR. AIH-tp1, autoimmune hepatitis time-point 1; AIH-tp2, autoimmune hepatitis time-point 2; CpGs, cytosine-phosphate-guanine dinucleotides; DMPs, differentially methylated probes; TSS, transcription start sites; IGR, intergenic regions; 5΄UTR, 5΄untranslated region; 3΄UTR, 3΄untranslated region; hyper, Hypermethylated; Hypo, hypomethylated.

Although GOmeth did not reveal any enriched pathway, DAVID could categorize 376/576 genes in 12 clusters and 38 subgroups (Supplementary Table 2). The main functional annotations over-represented were those classified as “nucleotide-binding kinases”, “metal-binding proteins”, “phospholipid-metabolism”, “motor-proteins”, “membrane-proteins” (p<0.05; Supplementary Table 2). Of note, the annotation cluster “immunity”, comprised of genes most of which were hypermethylated in AIH-tp2 (Supplementary Table 3).

To go further in the analysis of immunosuppression on DNA methylation in CD4(+) lymphocytes, the 14 DMRs between AIH-tp1 and AIH-tp2 (Supplementary Table 4) as well as the top 25 differentially methylated genes (among genes with 2 DMPs) were explored (Table 2). DMRs surrounding promoter regions were predominantly retrieved in 9/14 (64.3%) of AIH-tp2 compared to AIH-tp1. Among the key immune genes differentially methylated between AIH-tp2 and AIH-tp1, the activation marker CD86, the miRNA processing enzyme DROSHA and lncRNAs (LINC00211 and LINC01140) were hypermethylated in AIH-tp2 (p<0.05; Table 2).

**Table 2:**
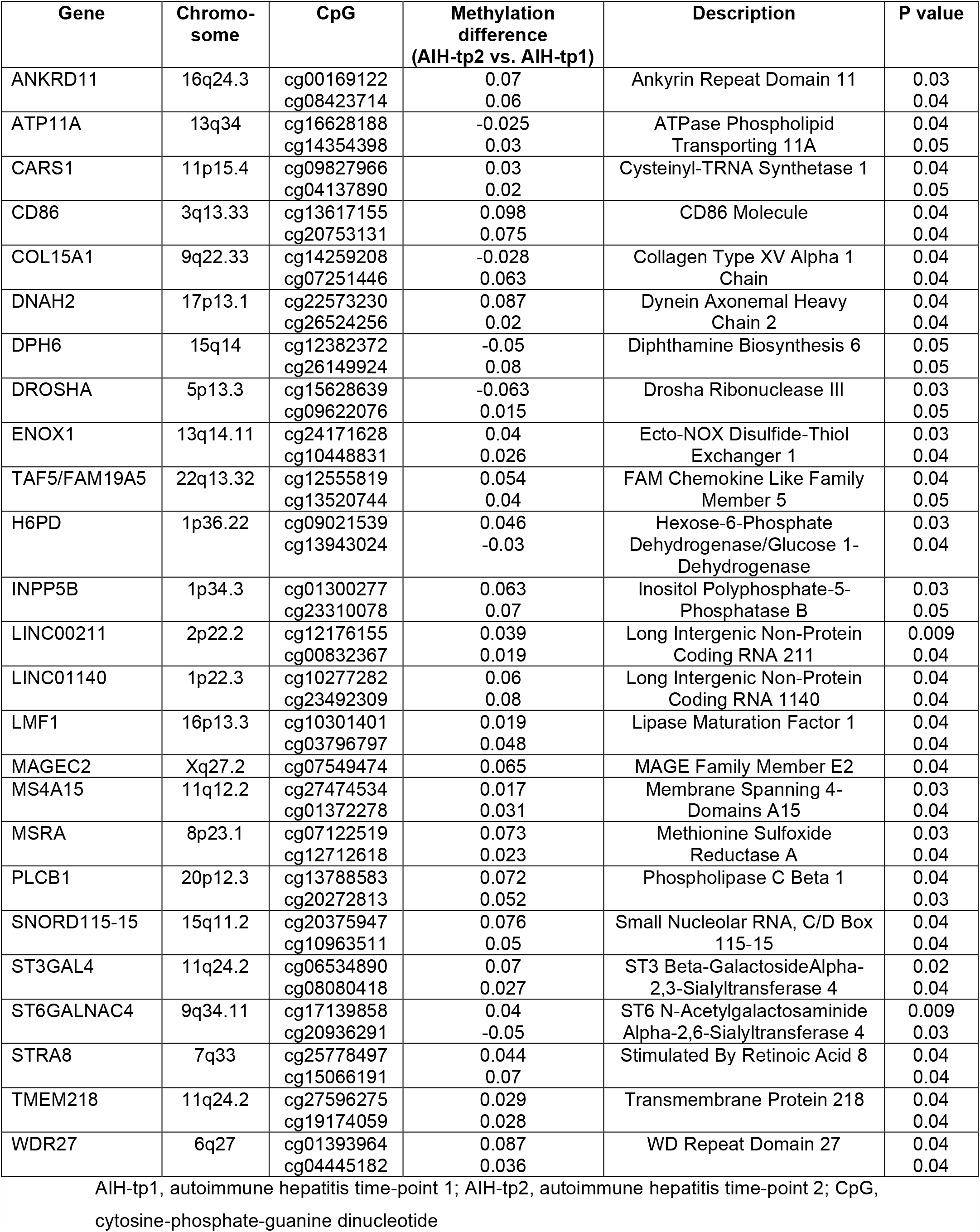
Top 25 differentially methylated genes between AIH-tp2 and AIH-tp1 patients.

### 5^m^C/5^hm^C staining in liver sections

Finally, representative intense and diffuse nuclear 5^hm^C immunohistochemical staining in the majority of the lymphocytes infiltrating the portal tract of AIH-tp1 cases is shown in Figure 6A. In addition, strong nuclear immunoreaction was observed in the limiting plate hepatocytes and bile duct epithelial cells of AIH-tp1 cases (Supplementary Table 5, Figure 6A). On the contrary, there was absence of positive lymphocytes in the portal tract of control cases, which also showed lack of immunoreactivity in periportal hepatocytes, and weak immunostaining of few bile duct epithelial cells (Supplementary Table 5, Figure 6B).

**Figure 6:**
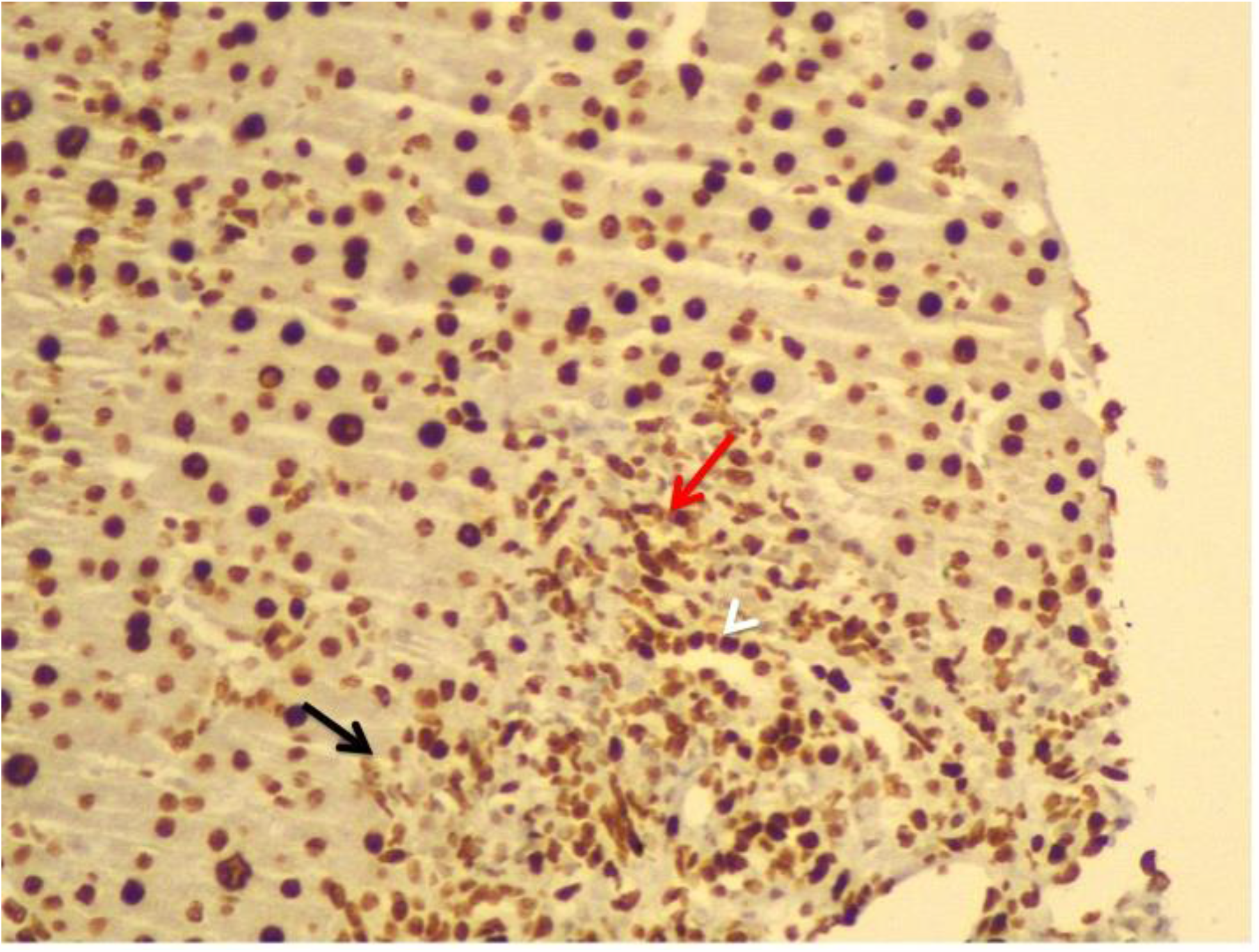
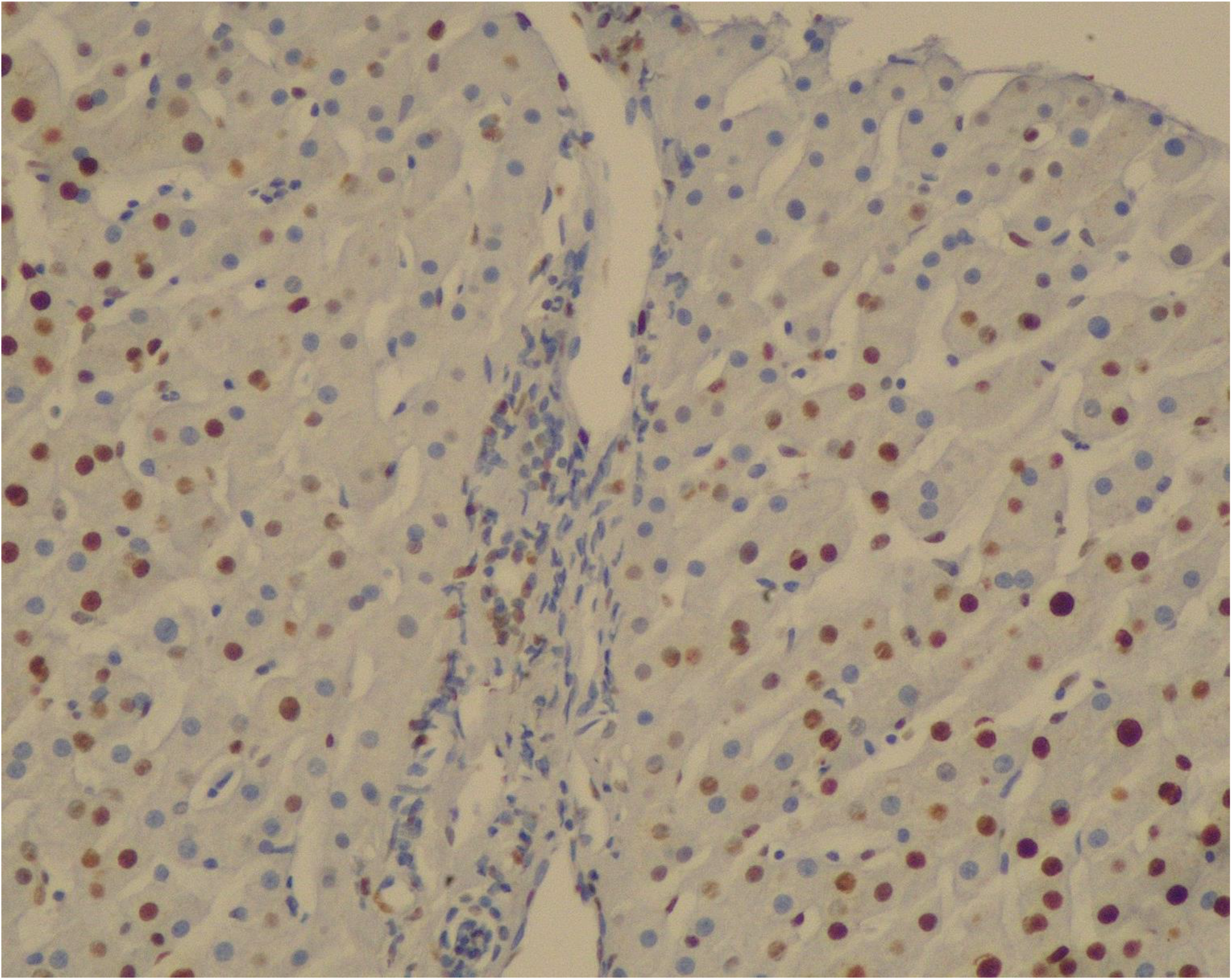
5^hm^C immunostaining, original magnification X200. **A)** AIH-tp1: 5^hm^C immunostaining of liver tissue sections shows strong nuclear positivity in the majority of the lymphocytes infiltrating the portal tract (red arrow). Portal inflammatory infiltrate disrupts limiting plate and surrounds individual hepatocytes. The latter show nuclear immunoreaction as well (black arrow). The bile duct epithelial cells show strong immunoexpression (arrowlead). **B)** Controls: 5^hm^C staining shows absence of positive lymphocytes in the portal tract. Note the absence of immunoreactivity in periportal hepatocytes. In addition, a few bile duct epithelial cells show a weaker positive immunoreaction than in AIH. 5^hm^C, 5-hydroxymethylcytosine; AIH-tp1, autoimmune hepatitis time-point 1.

Hepatocytes of both AIH-tp1 and control cases showed similar nuclear 5^hm^C and 5^m^C immunostaining (Supplementary Table 5 and 6, respectively). 5^m^C staining of liver infiltrating lymphocytes showed no differences in localization and intensity between patients and controls (Supplementary Table 6).

5^m^C and 5^hm^C immunostaining of Kupffer cells showed slightly reduced scores between AIH and HC (Supplementary Tables 5 and 6). However, the number of cases studied is small to allow further interpretation.

## Discussion

To the best of our knowledge, this is the first study to evaluate the DNA methylation status in liver sections and peripheral B- and T-cells from AIH patients. The following points arise from the present investigation: first, altered TET1 and DNMT3A expression characterizes both CD19(+) and CD4(+)-lymphocytes from AIH-tp1 patients compared to HC and PBC; second, after induction of remission, DNMT3A expression is decreased; third, in AIH-tp1 patients, DNMT3A was negatively correlated with IgG, while the opposite was observed in AIH-tp2 patients; forth, although changes in DNMT3A and TET1 expression were not associated with global and major 5^m^C/5^hm^C changes, using an EWAS approach we found however, differences of DNA methylation of specific genes in CD4(+)-lymphocytes from AIH-tp1 patients compared to HC and AIH-tp2 patients; and fifth, we observed strong nuclear 5^hm^C staining at the histological level of the periportal infiltrating lymphocytes in AIH-tp1 compared to controls. Taken together, these findings suggest that epigenetic modifications may play an important role in AIH pathogenesis and therapeutic response.

TETs are the main enzymes involved in active DNA demethylation associated with the modification and removal of 5^m^C. In mice, it has been shown that TETs play important role in mature B-cell antibody production, but also in facilitating the in-vitro differentiation of naïve CD4(+)-cells to T-regulatory cells (T-regs) by demethylating Foxp3 enhancer CNS2, while in-vivo seem to stabilize the expression of Foxp3 in T-regs (33). Therefore, TET deficient phenotypes are characterized by reduced class switch recombination capacity and decreased Foxp3 stability (33). Our findings of decreased transcriptional expression of TET1 in CD19(+)- and CD4(+)-lymphocytes from AIH patients with active disease may reflect the abovementioned immune dysregulation.

In AIH-tp1, DNMT3A expression in CD19(+)- and CD4(+)-lymphocytes was increased compared to PBC. This finding together with the increased DNMT1 and TET3 levels, which characterized CD19(+)-cells of PBC compared to HC, points to a different epigenetic profile between the two diseases. In addition, increased DNMT3A and DNMT1 transcriptional levels have been reported in CD4(+)-lymphocytes from patients with either clinically or serologically active systemic lupus erythematosus (SLE) (34). In SLE, both methyltransferases were inversely correlated with markers of disease activity such as, C3 complement component and anti-dsDNA antibody (34). Accordingly, we found that DNMT3A levels in active AIH were negatively associated with IgG, a serological marker of AIH activity. This correlation between DNMT3A mRNA levels and IgG may reflect a DNMT3A dependent plasma cell dysregulation, which increases autoantibody production as indicated by the elevated IgG values (35).

Corticosteroids administration has been reported to alter DNMT1 expression in PBMCs from SLE patients, while MMF has also been shown to induce epigenetic changes (36,37). This is in accordance with our findings, as immunosuppression with MMF seems to decrease DNMT3A expression in both CD19(+)- and CD4(+)-lymphocytes. The decrease of DNMT3A in responders compared to active AIH together with its positive correlation with IgG in patients at remission suggests that immunosuppression probably restores the epigenetic deregulations at least in B-cells.

EWAS in CD4(+)-cells showed methylation alterations of specific genes. Actually, DMRs analysis between AIH-tp1 and HC revealed that most DMRs located on gene promoters indicating their potential effect on the expression of implicated genes. However, recent studies have shown that the hyper/hypomethylation of the regions downstream of promoter-TSS are also highly involved in the regulation of gene expression (38).

Methylation changes in CD4(+)-cells affected, between others, HLA-DP genes. The association of AIH with the HLA class-II alleles was retrieved from early studies, and confirmed from recent genome wide association studies (GWAS) (7,8). In this context, HLA-DP polymorphisms can modulate interactions with the invariant chain chaperone, resulting in presentation of both exogenous and endogenous antigens in CD4(+)-cells (39). In addition, ZFP57 gene was found differentially methylated. ZFP57 belongs to the Methyl-CpG Binding Zinc Finger Proteins (a large family of methyl-binding proteins) and is associated with several cellular processes including regulation of gene expression, genomic imprinting, cell signalling and transcriptional repression (40).

LINC02571 gene was also found hypermethylated. Recent evidence indicates that lncRNAs play important roles in controlling the development of diverse immune cells and the mechanisms of immune cell activation (41). Of note, another gene located on MHC class-III, the TNF gene promoter, was found hypomethylated. This finding keeps up with a recent study of Bovensiepen et al (42), who found that TNF gene expression in liver-infiltrating lymphocytes was strongly upregulated and that the proportion of TNF-producing CD4(+)-cells was elevated both in blood and in the liver of AIH patients compared to HC. Among the rest of the genes found hypo/hypermethylated in AIH-tp1, RhoH (promoter hypermethylation) belongs to the Rho family of small GTPases and plays an important role in positive and negative thymic selection, in the functional differentiation of T-cells and in T-cell activation through TCR signalling (43). Interestingly, most of the genes (88%) of CD4(+)-cells were hypermethylated in AIH-tp2 patients, suggesting a shift in methylation profile after achievement of remission, probably towards a less activated state (37). Furthermore, some of these genes such as, ANXA1 and PYCARD are known to play diverse roles in immune responses, especially in the activation of inflammasome (44).

Among the top differentially methylated genes, CD86 encodes CD86/B7.2 molecule, which is a central costimulatory molecule mainly expressed on antigen presenting cells. However, recently it has been shown that CD86 is also expressed in humans on CD4(+)-cells in response to activation, suggesting a functional role of B7 molecules in the regulation of a T-cell response (45). The hypermethylation of CD86 gene in AIH at remission could signify a central role of CD86 molecule in “switch-off” of the immunological response in AIH. Interestingly, SorCS1 promoter was found hypomethylated in AIH at remission. This is in line with the role of the family of Vps10p receptors in the regulation of production and exocytosis of pro-inflammatory cytokines as well as the immune functions of T and NK-cells during adaptive immune responses (46).

In order to explore “in situ’’ modifications of methylation, that might contribute to AIH molecular phenotype and might influence the methylation status of circulating immune cells, we assessed the 5^m^C and 5^hm^C protein expression in paraffin embedded liver sections. Interestingly, the findings confirmed the results of EWAS since the majority of the lymphocytes infiltrating the portal tract, and the surrounding individual periportal hepatocytes, in AIH-tp1 liver sections, were hypomethylated (stained strongly for 5^hm^C) compared to HC. Of note, periportal hepatocytes and biliary duct epithelial cells seemed to follow the same pattern of hypomethylation, pointing towards an altered methylation milieu in the liver of AIH patients. To our knowledge, this is the first report of the 5^hm^C and 5^m^C liver tissue-mapping of AIH. These findings merit further investigation in large scale studies including a large number of liver biopsies, in order to correlate the specific cellular hypomethylation with the pathogenesis and/or the progression of the disease.

Our study has some limitations as the number of patients and controls was limited. However, the fact that we studied epigenetic modifications in pure peripheral B- and T-cells but also in liver sections, increases the reliability of our findings, as epigenetic changes are cell-type specific and studies on mixed cell populations may lead to ambiguous results.

In conclusion, we showed for the first time, that altered expression of DNMT3A and TET1, as well as altered DNA methylation of specific genes characterize immune cells in periphery and at the histological level of AIH patients, supporting the implication of epigenetic modifications in disease pathogenesis. Notably, epigenetic modifications were associated with disease activity and modified by immunosuppression. These findings open new insights in understanding of disease pathophysiology and may lead to novel therapeutic interventions.

## Data Availability

The data used to support the findings of this study are included within the article.

## Acknowledgements

This research project was supported in part by a research grant from the Hellenic Association for the Study of the Liver (HASL) and the Research Committee of the University of Thessaly (No. 2466). P. Arvaniti was also supported by a research grant for rare diseases by the Federation for the Development of Internal Medicine in Europe (FDIME).

## Abbreviations

5^hm^C: 5-hydroxymethyl-cytosine
5^m^C: 5-methyl-cytosine
AIH: Autoimmune hepatitis
ALT: Alanine aminotransferase
AMA: Antimitochondrial autoantibodies
ANA: Antinuclear autoantibodies
AST: Aspartate aminotransferase
DAVID: Database for Annotation, Visualization and Integrated Discovery
DMPs: Differentially methylated probes
DMRs: Differentially methylated regions
DNMTs: DNA methyl-transferases
EWAS: Epigenome wide association studies
GAPDH: Glyceraldehyde-3-Phosphate Dehydrogenase
GWAS: Genome wide association studies
HC: Health controls
HLA: Human leucocyte antigen
MHC: Major Histocompatibility complex
miRNAs: micro RNAs
MMF: Mycophenolate mofetil
NASH: Non alcoholic steatohepatitis
PBC: Primary biliary cholangitis
PBMCs: Peripheral blood mononuclear cells
SjS: Sjögren’s syndrome
SLA: Soluble liver antigen
SLE: Systemic lupus erythematosus
SMA: Smooth muscle cell antibodies
TETs: Ten-eleven translocation deoxygenases
TSS: Transcription start sites
tp1: time point 1
tp2: time point 2
Tregs: T regulatory
TSS: Transcription start site
UTR: Untranslated region

