## Supplemental figures for "Altered DNA methylation pattern characterizes the peripheral immune cells of patients with autoimmune hepatitis"

**Legend to supplementary figures**

**Supplementary Figure 1.** Purity of the isolated CD19(+) B and CD4(+) T-cells was assessed by flow cytometry using PE-Cy5-anti-CD19 and FITC-anti-CD3/PE-anti-CD8 antibody, respectively. **A)** Cluster of CD19(+) B-cells. **B)** Cluster of CD4(+) T-cells. **C)** Incubation of CD19 (+)-lymphocytes with PE-Cy5-anti-CD19 antibody showing a pure population of CD19(+) cells (purity 97.5%), while incubation with FITC-anti-CD3 shows no T-cell population. **D)** Incubation of CD4(+)-lymphocytes with FITC-anti-CD3/PE-anti-CD8 antibody showing a pure population of CD4(+) T-cells (CD3(+)/CD8(-) cells) (purity 97.8%).

**Supplementary Figure 2.** PBC patients had increased DNMT1 (**A**) and TET3 (**B**) levels in CD19(+) B-cells compared to HC (p=0.02 and p=0.005, respectively). AIH-tp1, autoimmune hepatitis time point 1 (at diagnosis, n=10); PBC, primary biliary cholangitis (n=9); HC, healthy controls (n=10); DNMT1, DNA methyltransferase 1; TET3, Ten-eleven translocation methylcytosine dioxygenase 3.

**Supplementary Figure 3.** In AIH-tp2, DNMT3A levels returned to normal, since they did not differ from HC or PBC, either in CD19(+) B (**A**) or in CD4(+) T-cells (**B**). AIH-tp2, autoimmune hepatitis time point 2 (at remission, n=8); PBC, primary biliary cholangitis (n=9); HC, healthy controls (n=10); DNMT3A, DNA methyltransferase 3A.

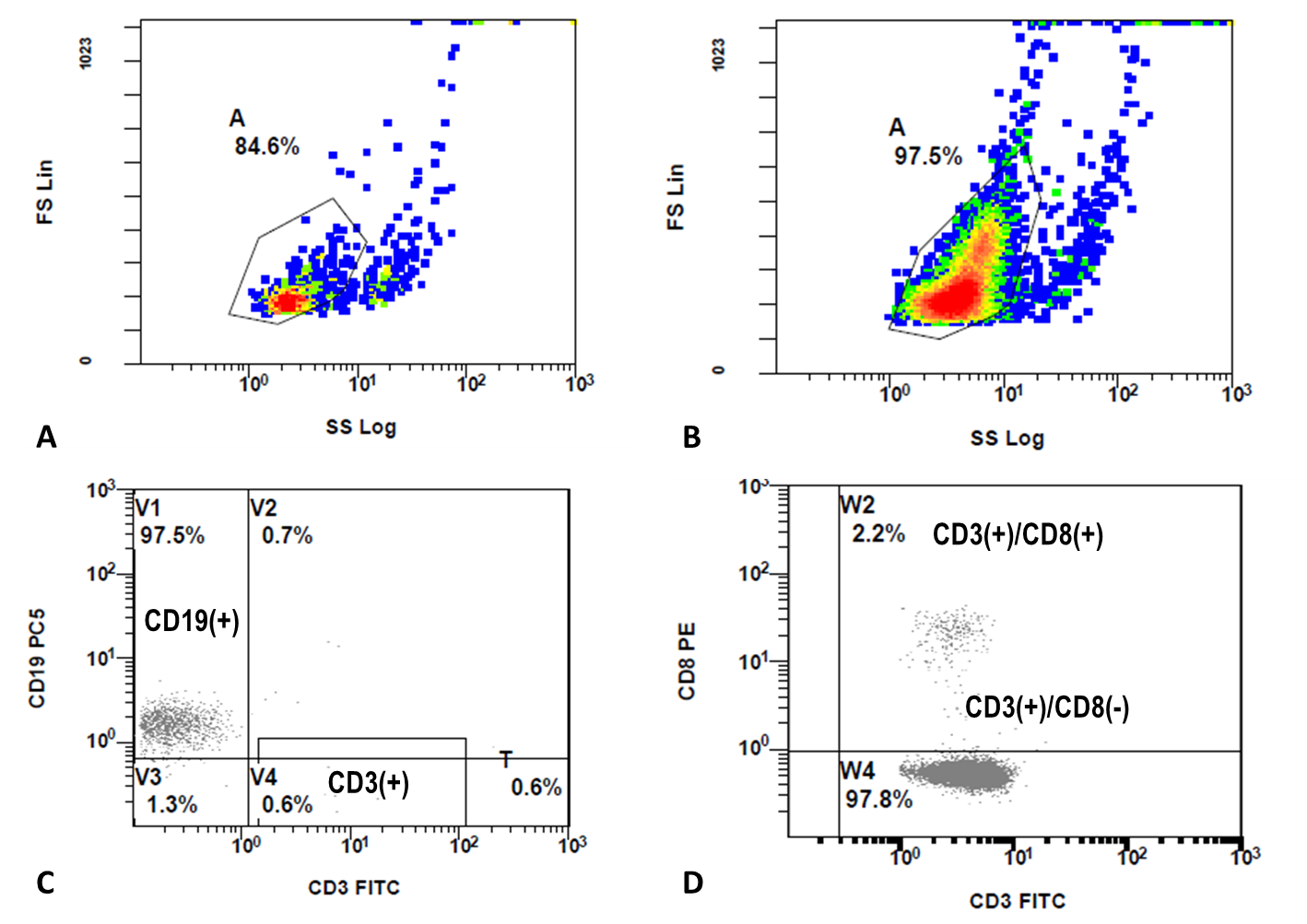

Supplementary Figure 1.

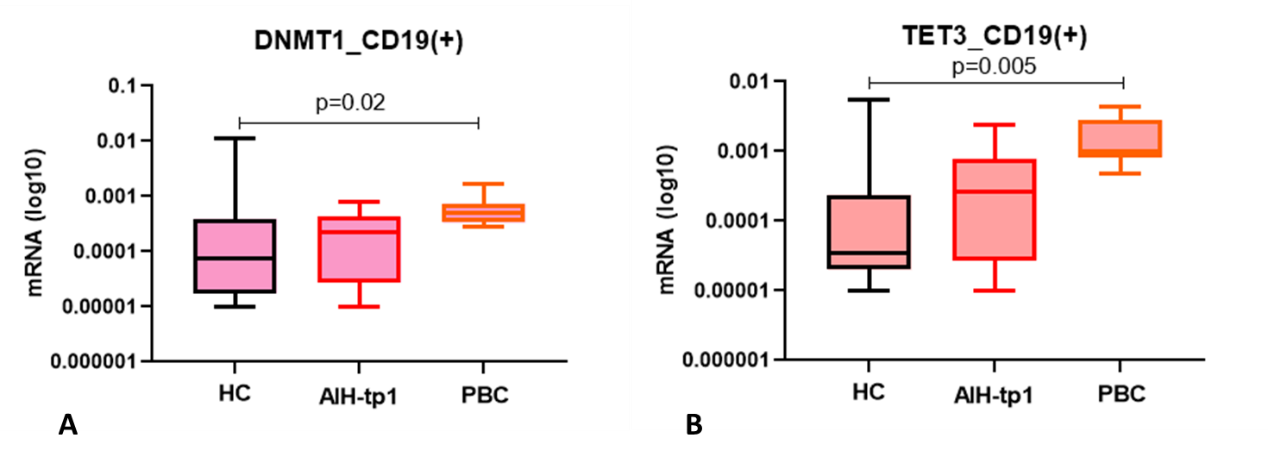

Supplementary Figure 2.

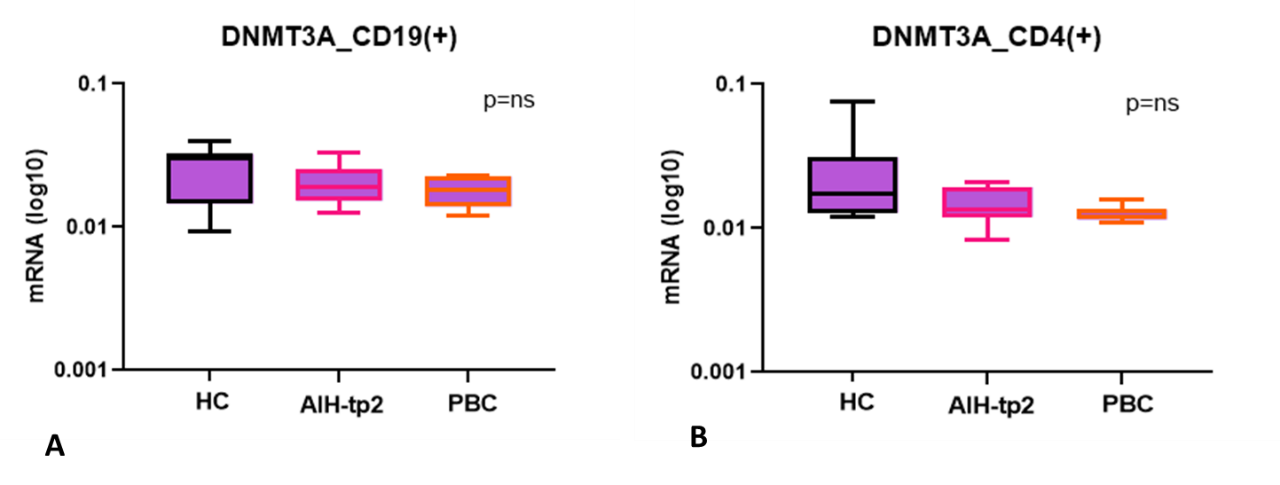

Supplementary Figure 3.
