## Supplemental Tables and Text for "Altered DNA methylation pattern characterizes the peripheral immune cells of patients with autoimmune hepatitis"

Supplementary Table 1: Clinical, biochemical, serological and histological characteristics from patients and HC.

| **Patients (n=37)** | **AIH tp1**  **(n=10)** | **AIH tp2**  **(n=8)** | **PBC**  **(n=9)** | **HC**  **(n=10)** |
| --- | --- | --- | --- | --- |
| Age (years) | 54 (33-77) | 54 (33-78) | 58 (37-76) | 48 (29-78) |
| Gender (male/female) | 2/8 | 2/6 | 0/9 | 3/7 |
| Disease duration (months) | 29±42 | 42±44 | 53±57 | - |
| Cirrhosis (yes/no) | 0/10 | 0/8 | 0/9 | - |
| Treatment (yes/no)  Prednisolone (yes/no)  MMF (yes/no) | 0/10 | 8/0  6/2  8/0 | 0/9  0/9  0/9 | -  -  - |
| Duration of treatment (months) | - | 6 (3-13) | - | - |
| AST (<40 IU/L)  ALT (<40IU/L)  Bilirubin (0.1-1mg/dl)  γGT (<45U/L)  ALP (<120U/L)*  IgG (<1650mg/dl)  IgM (64-249mg/dl) | 184±166  250±191  3.3±3.7  111±92  0.8±0.2  2244±449  181±109 | 24.5±6  18±6  0.8±0.6  23.7±25  0.5±0.13  1054±325  74.6±28.6 | 38±16  52±40  0.8±0.6  131±116  2.2±3.1  1333±191  324±228 | 20±3,4  20±7,6  0,6±0,32  20±11  0.6±0.1  866±135  115±38 |
| ANA (positive/negative)  SMA (positive/negative)  AMA (positive/negative)  SLA/LP (positive/negative) | 3/7  10/10  0/9  1/9 | 1/7  7/1  0/8  0/8 | 1/8  0/9  9/9  0/9 | 0/10  0/10  0/10  0/10 |
| Histology  Grading (minimal-mild/  moderate-severe)  Staging (minimal/mild-moderate/ severe-cirrhosis)  Ludwig (stage I/II/III/IV) | n=9  2/7  8/1  - | n=3  3/0  3/0  - | n=9  -  -  3/4/2/0 | - |
| Revised AIΗ score**  Simplified AIH score*** | 15.6±2.9  7±1 | 18±3.4  7±1 | -  - | -  - |

AIHtp1, autoimmune hepatitis timepoint 1 (at diagnosis); AIHtp2, autoimmune hepatitis timepoint 2 (at remission); PBC, primary biliary cholangitis; HC, healthy controls; AST: aspartate aminotransferase; ALT, alanine aminotrasferase; γGT, gamma glutamyl transferase; ALP, alkaline phosphatase; ANA, antinuclear antibodies; AMA, antimitochondrial antibodies; SMA, smooth muscle cells-antibodies; SLA/LP, soluble liver antigen/liver pancreas; *values are expressed as ratio to the upper limit of normal; **>15 definite ΑIΗ pre-treatment, >17 definite AIH post treatment; ***≥7 definite ΑIΗ

Supplementary Table 2. [DAVID (Database for Annotation, Visualization and Integrated Discovery) functional annotation bioinformatics microarray analysis](https://david.ncifcrf.gov/) could categorize 376 (out of 576) genes in 12 clusters and 38 subgroups.

| Annotation Cluster 1 | Enrichment Score: 3.98 |  |  |
| --- | --- | --- | --- |
| Category | Term | Genes | P-Value |
| UP_KEYWORDS | ATP-binding | CARS, ATP8A2, MAST3, PRKAG2, STK19, SMC4, AACS, RPS6KA2, AKT3, RUVBL1, TLK1, EPHB3, MAGI1, UPF1, RPS6KL1, CAMK1D, ACSL1, STARD9, MATK, UBE2E2, CSNK1E, ATP11A, KIF22, COASY, ATAD3C, DDX19B, OBSCN, DGKH, IPPK, CAMK2B, PFKFB4, DNAH1, DNAH2, ADCY3, AK4, NUAK1, ERBB2, MKNK2, MOV10L1, DPH6, VARS2, MARK3, TRAP1, DNAH17, CHKA, DMPK, TNK2, GATB, UBE2G1, TRPV1, CLK3, ADCY10, SBK1, EHD4, STK24, WNK2, NEK10, MAP3K10 | 3,54E+10 |
| UP_KEYWORDS | Kinase | IPPK, CAMK2B, PFKFB4, MAST3, PRKAG2, AKAP7, AK4, STK19, SGMS2, NUAK1, RPS6KA2, ERBB2, AKT3, MKNK2, TLK1, MARK3, EPHB3, RPS6KL1, CAMK1D, CHKA, DMPK, TNK2, MATK, CSNK1E, COASY, CLK3, OBSCN, SBK1, PRKAR1B, STK24, WNK2, NEK10, MAP3K10, ALPK3, DGKH | 1,67E+12 |
| UP_KEYWORDS | Nucleotide-binding | CARS, RAB3B, ATP8A2, MAST3, PRKAG2, STK19, SMC4, AACS, SEPT6, RPS6KA2, AKT3, RUVBL1, TLK1, EPHB3, MAGI1, UPF1, RPS6KL1, CAMK1D, ACSL1, STARD9, MATK, UBE2E2, CSNK1E, ATP11A, KIF22, COASY, ATAD3C, DDX19B, OBSCN, PRKAR1B, CNGB3, PDE5A, SLC27A3, DGKH, IPPK, CAMK2B, PFKFB4, DNAH1, DNAH2, ADCY3, AKAP7, AK4, FHIT, NUAK1, ERBB2, MKNK2, MOV10L1, DPH6, VARS2, MARK3, TRAP1, DNAH17, CHKA, DMPK, TNK2, GATB, UBE2G1, TRPV1, CLK3, ADCY10, SBK1, EHD4, STK24, WNK2, NEK10, MAP3K10, TUBA8 | 1,97E+11 |
| Annotation Cluster 2 | Enrichment Score: 1.69 |  |  |
| Category | Term | Genes | P-Value |
| UP_KEYWORDS | Metal-binding | CARS, ZNF296, ATP8A2, GALNT18, ITSN1, LOXL3, EHMT1, LOXL4, ZBTB22, NR3C1, ADARB2, SLC8A1, ALKBH3, ZFYVE26, STS, ZMIZ1, TRIM29, RPS6KA2, FNTB, ENPP2, TRIM26, TGM6, BSN, VPS18, RNF43, KDM2B, ZNF281, NSMCE2, ATP11A, OBSCN, KAT6A, PGR, DGKH, ZNF792, MTMR3, PDE1C, DTX1, RNF219, ZNF26, INPP5B, NUAK1, ADAMTS17, PRDM16, AMZ2, HIVEP3, ARSG, PLCG1, LHPP, ZSCAN18, ZNF660, ZFHX3, ACE, DMPK, ZBTB16, PHF12, TNK2, TRPV1, PHC3, CRNN, ADCY10, EHD4, COPS5, STK24, MAN2B2, OGDH, ALPI, GALNTL6, NRP2, LPCAT1, ZNF48, SGSH, SLC6A4, GLI2, ING5, MECOM, ZNF408, ADAMTS7, UPF1, KLF12, ARG2, ANXA1, TNNC2, MSL2, ZNF57, RCN2, RRM2B, PITPNM1, MMP15, PXDN, TAX1BP1, PDE5A, ZNF512, B4GALT5, MGRN1, HAAO, NR1I3, ADCY3, ZNF518A, ASAP2, ABLIM2, POLR2A, PLAGL1, MKNK2, DROSHA, METAP1D, KDM4B, KCNIP2, PML, SH3RF3, CYP2W1, ZNF74, NEK10, LNX1, ESYT2, TRIML1 | 7,33E+11 |
| UP_KEYWORDS | Zinc | CARS, ZNF296, EHMT1, ZBTB22, NR3C1, ADARB2, ZNF48, GLI2, ING5, ZFYVE26, MECOM, ZNF408, ZMIZ1, TRIM29, FNTB, ENPP2, TRIM26, BSN, ADAMTS7, VPS18, UPF1, RNF43, KLF12, KDM2B, ZNF281, NSMCE2, MSL2, SLC39A13, ZNF57, MMP15, KAT6A, TAX1BP1, PGR, ZNF512, DGKH, ZNF792, MTMR3, MGRN1, NR1I3, DTX1, RNF219, ZNF518A, ASAP2, ZNF26, ABLIM2, POLR2A, PLAGL1, MKNK2, ADAMTS17, PRDM16, HIVEP3, AMZ2, ZSCAN18, ZNF660, ZFHX3, KDM4B, ACE, ZBTB16, PHF12, PHC3, PML, SH3RF3, MAN2B2, ZNF74, LNX1, ALPI, TRIML1 | 0.07 |
| UP_KEYWORDS | Zinc-finger | ZNF296, ZBTB22, NR3C1, ZNF48, GLI2, ING5, ZFYVE26, MECOM, ZNF408, ZMIZ1, TRIM29, TRIM26, BSN, VPS18, UPF1, RNF43, KLF12, KDM2B, ZNF281, NSMCE2, MSL2, ZNF57, KAT6A, TAX1BP1, PGR, ZNF512, DGKH, ZNF792, MTMR3, MGRN1, NR1I3, DTX1, RNF219, ZNF518A, ZNF26, PLAGL1, PRDM16, HIVEP3, ZSCAN18, ZNF660, ZFHX3, KDM4B, ZBTB16, PHF12, PHC3, PML, SH3RF3, ZNF74, LNX1, TRIML1 | 0.15 |
| Annotation Cluster 3 | Enrichment Score: 1.12 |  |  |
| Category | Term | Genes | P-Value |
| UP_KEYWORDS | Phospholipid metabolism | PTPRN2, CHKA, LPCAT3, LPCAT1, AGPAT1 | 0.03 |
| UP_KEYWORDS | Phospholipid biosynthesis | CHKA, LPCAT3, LPCAT1, AGPAT1 | 0.09 |
| UP_KEYWORDS | Lipid biosynthesis | CHKA, LPCAT3, MOGAT1, LPCAT1, PRKAG2, AGPAT1, HACD2 | 0.16 |
| Annotation Cluster 4 | Enrichment Score: 0.97 |  |  |
| Category | Term | Genes | P-Value |
| UP_KEYWORDS | Dynein | DNAH1, DNAH2, DNAH17, DYNLRB1 | 0.04 |
| UP_KEYWORDS | Microtubule | DNAH1, DNAH2, DNAH17, TUBGCP3, MAPRE3, MAPT, DYNLRB1, KIF22, DISC1, CLASP2, TUBA8 | 0.13 |
| UP_KEYWORDS | Motor protein | DNAH1, DNAH2, DNAH17, STARD9, DYNLRB1, KIF22 | 0.21 |
| Annotation Cluster 5 | Enrichment Score: 0.82 |  |  |
| Category | Term | Genes | P-Value |
| UP_KEYWORDS | Membrane | ITSN1, EHMT1, LDLRAD3, PCMT1, STS, AKT3, MS4A15, MS4A13, PTGFRN, FAM19A5, SLC12A8, EPHB3, IER3, VPS18, UTS2R, SLC6A17, ACSL1, GPR37L1, ANK1, SCAMP2, TIAM1, ADGRB2, CCNY, PRKAR1B, KCNQ1, ATP6V0D1, LLGL1, MTMR3, TTYH3, SDC3, SLC5A1, HACD2, INPP5B, TRPM1, TSPAN9, AGMO, ST3GAL4, ST8SIA6, KCNN3, KCNN4, B4GALNT3, SNX8, GSDMC, TRAP1, ACE, PTPRN2, ICMT, DMPK, LRBA, BTBD11, MSRA, PEX5L, EHD4, WNK2, GALNTL6, SFXN5, CPEB4, GABRB3, CLIC6, OR11L1, NRP2, TENM4, MOGAT1, LPCAT1, SECTM1, TMEM51, SNX10, SIPA1L3, SLC6A4, PANX1, BBS9, CD38, RRP12, MUC15, DLGAP4, OR4D2, ANXA1, NFAM1, MYOF, ZDHHC13, EMP1, SLC39A13, SORCS1, IL17RC, BACE1, PSMA1, PITPNM1, MMP15, ARHGEF4, MAPT, MFGE8, PLCB1, KANK1, GPR45, TMPRSS6, ADCY3, AKAP7, SGMS2, PROKR1, CHRNE, C16ORF91, DISC1, LY6G6D, RHBG, EVC2, AP3D1, MPP5, CYP2W1, C6ORF47, DLC1, LPCAT3, ESYT2, RAMP1, RAB3B, CD86, ATP8A2, GALNT18, NCF4, ARHGAP35, SLC8A1, RUVBL1, ENPP2, SLC25A46, KCNH1, SH3GL1, MAGI1, RNF43, MATK, TTC7B, CDYL, ATP11A, TMEM130, SRCIN1, PROCR, MYADM, LY6D, PGR, TSNARE1, SLC27A3, WDFY4, DGKH, IFNAR1, FBN2, NDRG2, RTN4, DPP6, MARK3, LRRC4B, ANKRD29, OSBPL6, TNK2, TRPV1, MCC, LY75-CD302, SLC2A9, ADCY10, SDK1, G6PC2, DLG2, STK24, ATG16L1, NMUR1, ALPI, TUBA8, SPRED2, DGCR2, SUN3, AXIN1, NRG1, RANGAP1, DDX19B, TMEM214, RRM2B, TMEM218, CNGB3, SCN4A, ST6GALNAC4, SHANK2, B4GALT5, CNTNAP4, CAMK2B, MGRN1, RRBP1, ASAP2, MFF, AGPAT1, STRN3, LMF1, LRP6, NCLN, CLMN, GNG7, VPS53, ERBB2, BSND, CACNG1, SLC25A20, CLASP2, OPCML, MLXIP, KCNIP2, SEMA4F, BAIAP2, PML, SYT17, ENOX1 | 0.005 |
| UP_KEYWORDS | Glycoprotein | CD86, GALNT18, LOXL3, LOXL4, LDLRAD3, SLC8A1, STS, AKT3, ENPP2, PTGFRN, BSN, SLC12A8, EPHB3, IER3, KCNH1, UTS2R, RNF43, SLC6A17, ACSL1, GPR37L1, TMEM130, TANC2, PROCR, ADGRB2, CRISPLD2, KCNQ1, LY6D, IFNAR1, FBN2, LRPAP1, TTYH3, H6PD, NXPH4, SDC3, SLC5A1, HACD2, IGSF21, DPP6, TSPAN9, ADAMTS17, ST3GAL4, SERPINH1, ST8SIA6, ARSG, LRRC4B, ACE, PTPRN2, TGFB3, WNT3A, TRPV1, LY75-CD302, SLC2A9, SDK1, G6PC2, MAN2B2, NMUR1, ALPI, GALNTL6, FBN1, GABRB3, OR11L1, NRP2, TENM4, MOGAT1, SECTM1, SGSH, SLC6A4, PANX1, CD38, MUC15, DGCR2, ADAMTS7, OR4D2, NFAM1, EMP1, NRG1, SORCS1, IL17RC, BACE1, PSMA1, TMEM214, MMP15, PXDN, CNGB3, SCN4A, MAPT, MFGE8, ST6GALNAC4, SHANK2, B4GALT5, CNTNAP4, COL15A1, GPR45, TMPRSS6, SERPINC1, LAMA3, ADCY3, LRP6, PROKR1, NCLN, CHRNE, ERBB2, CACNG1, LY6G6D, RHBG, OPCML, EVC2, GDF11, COL22A1, SEMA4F, CYP2W1, FMOD | 0.37 |
| UP_KEYWORDS | Transmembrane | CD86, ATP8A2, GALNT18, ITSN1, EHMT1, LDLRAD3, SLC8A1, PCMT1, STS, ENPP2, MS4A15, MS4A13, PTGFRN, FAM19A5, SLC12A8, SLC25A46, EPHB3, IER3, KCNH1, UTS2R, RNF43, SLC6A17, ACSL1, GPR37L1, CDYL, ATP11A, TMEM130, SCAMP2, PROCR, ADGRB2, KCNQ1, MYADM, TSNARE1, SLC27A3, WDFY4, IFNAR1, FBN2, TTYH3, SDC3, SLC5A1, NDRG2, HACD2, RTN4, TRPM1, DPP6, TSPAN9, AGMO, ST3GAL4, ST8SIA6, KCNN3, KCNN4, B4GALNT3, LRRC4B, ANKRD29, ACE, PTPRN2, ICMT, DMPK, LRBA, BTBD11, TRPV1, LY75-CD302, SLC2A9, ADCY10, SDK1, G6PC2, NMUR1, ALPI, GALNTL6, SFXN5, TUBA8, GABRB3, CLIC6, OR11L1, NRP2, TENM4, MOGAT1, LPCAT1, TMEM51, SECTM1, SLC6A4, PANX1, CD38, MUC15, RRP12, DGCR2, OR4D2, SUN3, NFAM1, MYOF, EMP1, ZDHHC13, NRG1, SLC39A13, SORCS1, IL17RC, BACE1, PSMA1, RRM2B, TMEM214, MMP15, TMEM218, CNGB3, ARHGEF4, SCN4A, ST6GALNAC4, SHANK2, B4GALT5, CNTNAP4, GPR45, TMPRSS6, ADCY3, RRBP1, SGMS2, AGPAT1, MFF, LMF1, LRP6, PROKR1, NCLN, CLMN, C16ORF91, CHRNE, ERBB2, BSND, CACNG1, SLC25A20, LY6G6D, RHBG, EVC2, SEMA4F, C6ORF47, LPCAT3, ESYT2, RAMP1 | 0.50 |
| UP_KEYWORDS | Transmembrane helix | CD86, ATP8A2, GALNT18, ITSN1, EHMT1, LDLRAD3, SLC8A1, PCMT1, STS, ENPP2, MS4A15, MS4A13, PTGFRN, FAM19A5, SLC12A8, SLC25A46, EPHB3, IER3, KCNH1, UTS2R, RNF43, SLC6A17, ACSL1, GPR37L1, CDYL, ATP11A, TMEM130, SCAMP2, PROCR, ADGRB2, KCNQ1, MYADM, TSNARE1, SLC27A3, WDFY4, IFNAR1, FBN2, TTYH3, SDC3, SLC5A1, NDRG2, HACD2, RTN4, TRPM1, DPP6, TSPAN9, AGMO, ST3GAL4, ST8SIA6, KCNN3, KCNN4, B4GALNT3, LRRC4B, ANKRD29, ACE, PTPRN2, ICMT, DMPK, LRBA, BTBD11, TRPV1, LY75-CD302, SLC2A9, ADCY10, SDK1, G6PC2, NMUR1, GALNTL6, SFXN5, TUBA8, GABRB3, CLIC6, OR11L1, NRP2, TENM4, MOGAT1, LPCAT1, TMEM51, SECTM1, SLC6A4, PANX1, CD38, MUC15, RRP12, DGCR2, OR4D2, SUN3, NFAM1, MYOF, EMP1, ZDHHC13, NRG1, SLC39A13, SORCS1, IL17RC, BACE1, PSMA1, RRM2B, TMEM214, MMP15, TMEM218, CNGB3, ARHGEF4, SCN4A, ST6GALNAC4, SHANK2, B4GALT5, CNTNAP4, GPR45, TMPRSS6, ADCY3, RRBP1, SGMS2, AGPAT1, MFF, LMF1, LRP6, PROKR1, NCLN, CLMN, C16ORF91, CHRNE, ERBB2, BSND, CACNG1, SLC25A20, LY6G6D, RHBG, EVC2, SEMA4F, C6ORF47, LPCAT3, ESYT2, RAMP1 | 0.53 |
| Annotation Cluster 6 | Enrichment Score: 0.63 |  |  |
| Category | Term | Genes | P-Value |
| UP_KEYWORDS | Cell division | IST1, NSMCE2, WASL, NR3C1, SMC4, CDC20, STAG1, ZFYVE26, SEPT6, CCNY, RUVBL1, MAPRE3, CEP63, CLASP2 | 0.13 |
| UP_KEYWORDS | Mitosis | CDC20, STAG1, NSMCE2, RUVBL1, MAPRE3, WASL, CEP63, NR3C1, SMC4, CLASP2 | 0.17 |
| UP_KEYWORDS | Cell cycle | IST1, NSMCE2, WASL, FOXN3, NR3C1, SMC4, CDC20, STAG1, ZFYVE26, SEPT6, CCNY, RUVBL1, TLK1, MAPRE3, CEP63, CLASP2 | 0.57 |
| Annotation Cluster 7 | Enrichment Score: 0.51 |  |  |
| Category | Term | Genes | P-Value |
| UP_KEYWORDS | Transcription | SETD2, ZNF296, CALCOCO1, WASL, ZBTB22, NR3C1, ZNF48, ARHGAP35, GLI2, ING5, MECOM, ZNF408, ZMIZ1, RUVBL1, LRRFIP1, TEAD2, KLF12, DAXX, KDM2B, ZNF281, RFX2, CDYL, ZNF57, SND1, POU5F1, NCOR2, KAT6A, IRF2, PGR, ZNF512, MAML3, KANK2, KANK1, ZNF792, NR1I3, ZNF518A, MLLT1, PRDM11, FHIT, ZNF26, STRA8, POLR2A, PLAGL1, ERBB2, PRDM16, HIVEP3, DEDD, ZSCAN18, MLXIP, ZNF660, ZFHX3, KDM4B, ZBTB16, PHF12, FOXN3, PTPN14, PML, POLR3D, NFIC, ZNF74, TADA1, POLR3G, TCF4, GTF2IRD1, NOL11 | 0.16 |
| UP_KEYWORDS | Transcription regulation | SETD2, ZNF296, CALCOCO1, WASL, ZBTB22, NR3C1, ZNF48, ARHGAP35, GLI2, ING5, MECOM, ZNF408, ZMIZ1, RUVBL1, LRRFIP1, TEAD2, KLF12, DAXX, KDM2B, ZNF281, RFX2, CDYL, ZNF57, SND1, POU5F1, NCOR2, KAT6A, IRF2, PGR, ZNF512, MAML3, KANK2, KANK1, ZNF792, NR1I3, ZNF518A, MLLT1, PRDM11, FHIT, ZNF26, STRA8, PLAGL1, ERBB2, PRDM16, HIVEP3, DEDD, ZSCAN18, MLXIP, ZNF660, ZFHX3, KDM4B, ZBTB16, PHF12, FOXN3, PTPN14, PML, NFIC, ZNF74, TADA1, TCF4, GTF2IRD1, NOL11 | 0.22 |
| UP_KEYWORDS | DNA-binding | ZNF792, NANOGNB, NR1I3, ZNF518A, ZBTB22, NR3C1, ZNF26, ZNF48, ARHGAP35, GLI2, MECOM, POLR2A, ZNF408, PLAGL1, PCBP2, PRDM16, LRRFIP1, TEAD2, DEDD, ZSCAN18, MLXIP, ZNF660, KLF12, ZFHX3, ZBTB16, ZNF281, STRBP, H3F3A, RFX2, FOXN3, KIF22, ZNF57, PHC3, POU5F1, PML, NCOR2, NFIC, ZNF74, IRF2, TCF4, GTF2IRD1, PGR, ZNF512, AHDC1 | 0.82 |
| Annotation Cluster 8 | Enrichment Score: 0.45 |  |  |
| Category | Term | Genes | P-Value |
| UP_KEYWORDS | Centromere | DAXX, STAG1, SEPT6, CENPN, RANGAP1, CLASP2 | 0.22 |
| UP_KEYWORDS | Kinetochore | SEPT6, CENPN, RANGAP1, CLASP2 | 0.41 |
| UP_KEYWORDS | Chromosome | SETD2, DAXX, STAG1, SEPT6, NSMCE2, H3F3A, EHMT1, CENPN, RANGAP1, SMC4, CLASP2 | 0.46 |
| Annotation Cluster 9 | Enrichment Score: 0.33 |  |  |
| Category | Term | Genes | P-Value |
| UP_KEYWORDS | Voltage-gated channel | CLIC6, KCNIP2, KCNQ1, SCN4A, CACNG1, KCNH1 | 0.28 |
| UP_KEYWORDS | Potassium transport | KCNIP2, KCNQ1, SLC12A8, KCNH1 | 0.51 |
| UP_KEYWORDS | Potassium channel | KCNIP2, KCNQ1, KCNH1 | 0.52 |
| UP_KEYWORDS | Potassium | KCNIP2, KCNQ1, SLC12A8, KCNH1 | 0.59 |
| Annotation Cluster 10 | Enrichment Score: 0.28 |  |  |
| Category | Term | Genes | P-Value |
| UP_KEYWORDS | Metalloprotease | ACE, COPS5, MMP15, ADAMTS17, AMZ2, ADAMTS7 | 0.29 |
| UP_KEYWORDS | Protease | USP24, METAP1D, ACE, USP42, TMPRSS6, BACE1, DPP6, COPS5, PSMA1, MMP15, ADAMTS17, AMZ2, ADAMTS7 | 0.63 |
| UP_KEYWORDS | Zymogen | BACE1, TMPRSS6, MMP15, ADAMTS17, ADAMTS7 | 0.74 |
| Annotation Cluster 11 | Enrichment Score: 0.28 |  |  |
| Category | Term | Genes | P-Value |
| UP_KEYWORDS | Antiviral defense | ELMOD2, POLR3D, PCBP2, POLR3G, PML | 0.29 |
| UP_KEYWORDS | Innate immunity | PYCARD, CHID1, ANXA1, POLR3D, PCBP2, POLR3G, PML | 0.58 |
| UP_KEYWORDS | Immunity | PYCARD, CHID1, CD86, ANXA1, PSMA1, POLR3D, PCBP2, POLR3G, KCNN4, PML | 0.84 |
| Annotation Cluster 12 | Enrichment Score: 0.26 |  |  |
| Category | Term | Genes | P-Value |
| UP_KEYWORDS | Glycoprotein | CD86, GALNT18, LOXL3, LOXL4, LDLRAD3, SLC8A1, STS, AKT3, ENPP2, PTGFRN, BSN, SLC12A8, EPHB3, IER3, KCNH1, UTS2R, RNF43, SLC6A17, ACSL1, GPR37L1, TMEM130, TANC2, PROCR, ADGRB2, CRISPLD2, KCNQ1, LY6D, IFNAR1, FBN2, LRPAP1, TTYH3, H6PD, NXPH4, SDC3, SLC5A1, HACD2, IGSF21, DPP6, TSPAN9, ADAMTS17, ST3GAL4, SERPINH1, ST8SIA6, ARSG, LRRC4B, ACE, PTPRN2, TGFB3, WNT3A, TRPV1, LY75-CD302, SLC2A9, SDK1, G6PC2, MAN2B2, NMUR1, ALPI, GALNTL6, FBN1, GABRB3, OR11L1, NRP2, TENM4, MOGAT1, SECTM1, SGSH, SLC6A4, PANX1, CD38, MUC15, DGCR2, ADAMTS7, OR4D2, NFAM1, EMP1, NRG1, SORCS1, IL17RC, BACE1, PSMA1, TMEM214, MMP15, PXDN, CNGB3, SCN4A, MAPT, MFGE8, ST6GALNAC4, SHANK2, B4GALT5, CNTNAP4, COL15A1, GPR45, TMPRSS6, SERPINC1, LAMA3, ADCY3, LRP6, PROKR1, NCLN, CHRNE, ERBB2, CACNG1, LY6G6D, RHBG, OPCML, EVC2, GDF11, COL22A1, SEMA4F, CYP2W1, FMOD | 0.37 |
| UP_KEYWORDS | Disulfide bond | GABRB3, CD86, OR11L1, NRP2, TENM4, GALNT18, LOXL3, LOXL4, SECTM1, LDLRAD3, SLC6A4, SGSH, PCMT1, STS, AKT3, ENPP2, CD38, PTGFRN, EPHB3, DGCR2, ADAMTS7, OR4D2, UTS2R, RNF43, ANXA1, NFAM1, NRG1, GPR37L1, BACE1, PROCR, OBSCN, ADGRB2, PRKAR1B, CRISPLD2, MMP15, PXDN, LY6D, ALPK3, MAPT, MFGE8, NBL1, ST6GALNAC4, CNTNAP4, B4GALT5, IFNAR1, FBN2, COL15A1, TMPRSS6, SERPINC1, LAMA3, REG3G, SLC5A1, LYZL6, LRP6, PROKR1, IGSF21, DPP6, CHRNE, ERBB2, ADAMTS17, ST3GAL4, ST8SIA6, LY6G6D, LRRC4B, OPCML, GSDMC, GDF11, ACE, PSPN, TGFB3, WNT3A, CCL20, SEMA4F, LY75-CD302, SDK1, NMUR1, ALPI, GALNTL6, FMOD, RAMP1, FBN1 | 0.58 |
| UP_KEYWORDS | Signal | GABRB3, CD86, IPO11, NRP2, LPCAT1, LOXL3, LOXL4, SECTM1, LDLRAD3, CYB5D2, SLC8A1, SLC6A4, SGSH, STS, ENPP2, MUC15, PTGFRN, EPHB3, DGCR2, ADAMTS7, CHID1, RNF43, NFAM1, NRG1, SLC39A13, SORCS1, GPR37L1, TMEM130, ANK1, IL17RC, COASY, BACE1, PROCR, RCN2, ADGRB2, CRISPLD2, MMP15, PXDN, LY6D, SCN4A, MFGE8, NBL1, CNTNAP4, IFNAR1, FBN2, LRPAP1, COL15A1, TTYH3, H6PD, SERPINC1, NXPH4, LAMA3, REG3G, AGPAT1, ZNF26, LYZL6, LRP6, IGSF21, NCLN, TSPAN9, C16ORF91, CHRNE, ERBB2, ADAMTS17, ST3GAL4, SERPINH1, ARSG, LY6G6D, RHBG, MARK3, LRRC4B, OPCML, EVC2, MLXIP, GDF11, PTPRN2, ACE, PSPN, TGFB3, WNT3A, CCL20, COL22A1, SEMA4F, LY75-CD302, MSRA, SDK1, CYP2W1, MAN2B2, B9D1, ALPI, FMOD, RAMP1, FBN1 | 0.78 |

Supplementary Table 3. Analysis of the annotation cluster “immunity” from [DAVID (Database for Annotation, Visualization and Integrated Discovery).](https://david.ncifcrf.gov/)

| **Gene** | **Protein*** | **Main Pathways*** | **Methylation**  **AIH-tp2 vs AIH-tp1** | **Methylation difference**  **AIH-tp2 vs AIH-tp1** | **P-value** |
| --- | --- | --- | --- | --- | --- |
| ELMOD2 | Engulfment and motility Domain-Containing Protein 2 | Antiviral responses | hyper | 0.096 | 0.04 |
| POLR3D | Polymerase (RNA) III (DNA Directed) Polypeptide D | RNA Polymerase II transcription initiation and promoter clearance/ transcription of tRNA. | hyper | 0.03 | 0.04 |
| PCBP2 | Poly(RC) Binding Protein 2 | Induction of IFN-alpha/beta pathways | hyper | 0.037 | 0.03 |
| POLR3G | Polymerase (RNA) III (DNA Directed) Polypeptide G | Sensing and limiting infection by intracellular bacteria and DNA viruses. Acts as nuclear and cytosolic DNA sensor involved in innate immune response. | hypo | -0.02 | 0.04 |
| PML | Promyelocytic Leukemia protein | The protein encoded by this gene is a member of the tripartite motif (TRIM) family. Functions via its association with PML-nuclear bodies in tumor suppression, transcriptional regulation, apoptosis, senescence, DNA damage response, and viral defense mechanisms | hyper | 0.06 | 0.04 |
| PYCARD | Apoptosis-Associated Speck-Like Protein Containing A CARD | Functions as key mediator in apoptosis and inflammation through caspases | hyper | 0.06 | 0.04 |
| CHID1 | Chromodomain Helicase DNA Binding Protein 1 | Alters gene expression possibly by modification of chromatin structure thus altering access of the transcriptional apparatus to its chromosomal DNA template | hyper | 0.05 | 0.04 |
| ANXA1 | annexin A1 | Plays important role in the innate immune response as effector of glucocorticoid-mediated responses and regulator of the inflammatory process. Promotes chemotaxis of granulocytes and monocytes via activation of the formyl peptide receptors | hyper | 0.11 | 0.009 |
| PSMA1 | Proteasome 20S Subunit Alpha 1 | An essential function of a modified proteasome, the immunoproteasome, is the processing of class I MHC peptides. | hyper | 0.03 | 0.04 |
| KCNN4 | Potassium Calcium-Activated Channel Subfamily N Member 4 | Part of the predominant calcium-activated potassium channel in T-lymphocytes | hyper | 0.02 | 0.04 |
| CD86 | CD86 antigen | The protein that is a member of the immunoglobulin superfamily involved in the costimulatory signal essential for T-lymphocyte proliferation and interleukin-2 production, by binding CD28 or CTLA-4 | hyper | 0.07 | 0.04 |

### *Retrieved from GeneCards^®^: The Human Gene Database https://www.genecards.org/

Supplementary Table 4. Differentially methylated genes, corresponding to 14 DMRs (differentially methylated regions) between AIH-tp2 and AIH-tp1 patients.

| Gene | Chromo-some | Description | Location | Methylation difference (AIH-tp2 vs AIH-tp1) | P-value |
| --- | --- | --- | --- | --- | --- |
| ARRDC4 | 15 | Arrestin domain containing 4 | promoter-TSS | -0.72 | 0.001 |
| CSGALNACT1 | 8 | Chondroitin sulfate N-acetylgalactosaminyl-transferase 1 | promoter-TSS | 0.76 | 0.009 |
| DOCK1 | 10 | Dedicator of cytokinesis 1 | promoter-TSS | -0.87 | 0.007 |
| HLA-DMA | 6 | Major histocompatibility complex | promoter-TSS | 0.67 | 0.008 |
| ISM1 | 20 | Isthmin 1 | Intergenic | -0.72 | 0.001 |
| KIAA0040 | 1 | Uncharacterized Protein KIAA0040 | promoter-TSS | 0.75 | 0.01 |
| TRIM39-RPP21 | 6 | TRIM39-RPP21 | exon | 0.65 | 0.02 |
| HLA-DMB | 6 | HLA-DMB | intron | 0.67 | 0.008 |
| SUN1 | 7 | Sad1 And UNC84 Domain Containing 1 | intron | -0.67 | 0.03 |
| FAM53B-AS1 | 10 | FAM53B-AS1 | intron | 0.89 | 0.008 |
| S100A13 | 1 | S100 calcium binding protein A13 | promoter-TSS | 0.51 | 0.01 |
| SLFN12 | 17 | Schlafen family member 12 | promoter-TSS | 0.55 | 0.003 |
| SORCS1 | 10 | Sortilin related VPS10 domain containing receptor 1 | promoter-TSS | -0.78 | 0.02 |
| THRB | 3 | Thyroid hormone receptor beta | promoter-TSS | -0.66 | 0.006 |

AIH-tp1, autoimmune hepatitis time point 1 (at diagnosis, n=10); AIH-tp2, autoimmune hepatitis time point 2 (at remission, n=5); TSS: transcription start sites

Supplementary Table 5. Evaluation of 5^hm^C immunostaining in liver tissue sections

| Sample | Hepatocytes | Lymphocytes | BECs | Kupffer | Periportal lymphocytes vs rest | Liver histology |
| --- | --- | --- | --- | --- | --- | --- |
| AIH-tp1-1 | +3 | +3 | +3 | +1 | YES | interface+2/portal+3 |
| AIH-tp1-2 | +3 | +3 | +3 | +1 | YES | interface+3/portal+4 |
| AIH-tp1-3 | +3 | +3 | +3 | +2 | YES | interface+3/portal+2 |
| AIH-tp1-4 | +3 | +3 | +3 | +3 | NO | interface+4/portal+2 |
| AIH-tp1-5 | +3 | +3 | +3 | +2 | YES | interface+3/portal+3 |
| AIH-tp1-6 | +2 | +2 | +2 | +1 | YES | interface+4/portal+4 |
| CONTROL-1 | +3 | +3 | +3 | +1 | NO |  |
| CONTROL-2 | +2 | 0 | 0 | 0 | NO |  |
| CONTROL-3 | +3 | +2 | +3 | +2 | NO |  |
| CONTROL-4 | +3 | 0 | +1 | +1 | NO |  |
| CONTROL-5 | +3 | +2 | +2 | +2 | NO |  |
| CONTORL-6 | +3 | +2 | +2 | +2 | NO |  |
| CONTROL-7 | +3 | +2 | +2 | +2 | NO |  |

Immunostaining was assessed in hepatocytes, lymphocytes of the portal tract and lymphocytes invading the limiting plate (periportal lymphocytes) as well as in biliary duct epithelial cells (BECs) and Kupffer cells. The extent of staining was scored according to its amount and intensity, using a 4-point scoring system, as follows: 0 = no staining; 1 = positive nuclear staining in less than 20% of cells; 2 = 21-50% of positive cells; and 3 = positive nuclear staining in more than 50% of cells.

AIH-tp1, autoimmune hepatitis time point 1 (at diagnosis); CONTROL, healthy controls

Supplementary Table 6. Evaluation of 5^m^C immunostaining in liver tissue sections

| Sample | Hepatocytes | Lymphocytes | BECs | Kupffer | Periportal lymphocytes vs rest | Liver Histology |
| --- | --- | --- | --- | --- | --- | --- |
| AIH-tp1-1 | +2 | +3 | +1 | +1 | NO | interface+2/portal+3 |
| AIH-tp1-2 | +3 | +3 | +2 | +2 | NO | interface+3/portal+4 |
| AIH-tp1-3 | +3 | +3 | +3 | +2 | NO | interface+3/portal+2 |
| AIH-tp1-4 | +3 | +3 | +3 | +3 | NO | interface+4/portal+2 |
| AIH-tp1-5 | +3 | +3 | +3 | +2 | NO | interface+3/portal+3 |
| AIH-tp1-6 | +2 | +2 | +1 | +1 | NO | interface+4/portal+4 |
| CONTROL-1 | +3 | +2 | +1 | +1 | NO |  |
| CONTROL-2 | +3 | +2 | +1 | +1 | NO |  |
| CONTROL-3 | +3 | +3 | +3 | +3 | NO |  |
| CONTROL-4 | +3 | +3 | +3 | +3 | NO |  |
| CONTROL-5 | +3 | +2 | +2 | +2 | NO |  |
| CONTORL-6 | +3 | +3 | +3 | +2 | NO |  |
| CONTROL-7 | +3 | +3 | +3 | 0 | NO |  |

Immunostaining was assessed in hepatocytes, lymphocytes of the portal tract and lymphocytes invading the limiting plate (periportal lymphocytes) as well as in biliary duct epithelial cells (BECs) and Kupffer cells. The extent of staining was scored according to its amount and intensity, using a 4-point scoring system, as follows: 0 = no staining; 1 = positive nuclear staining in less than 20% of cells; 2 = 21-50% of positive cells; and 3 = positive nuclear staining in more than 50% of cells.

AIH-tp1, autoimmune hepatitis time point 1 (at diagnosis); CONTROL, healthy controls

*Patients and methods*

*Determination of 5^m^C and 5^hm^C DNA levels*

Briefly, 100ng of DNA were bound in duplicates to strip-wells with high DNA affinity, the methylated and hydroxymethylated fraction was detected by 5^m^C and 5^hm^C antibody-based detection complex in one-step manner and quantified colorimetrically at 450nm in a microplate spectrophotometer (Dynex OpsysMR^TM^ Microplate Reader, Aspect Scientific Ltd, UK). The percentage of 5^m^C and 5^hm^C in the cytosine content was proportional to the optical density (OD) measured and was calculated by generating a logarithmic standard curve based on positive, negative controls and seven standard samples supplied by the manufacturer.

EWAS

In brief, the DNA was deaminated with the EZ-96 DNA Methylation Kit (Zymo Research) according to Illumina’s recommended protocol. Bisulfite conversion was controlled by qPCR. One assay targeting a methylated region of DNAJC15 and two assays targeting the GNAS locus (one assay for the unmethylated allele and one for the methylated one) were used for quality control. Deaminated DNA derived from blood was amplified in parallel and served as positive control. A sample passed the quality control when the received CT-value either for the two GNAS loci or the DNAJC15 locus reaches the threshold not later than 5 cycles compared to the positive control.
